# A germline and somatic mutation sorting (GeMSort) algorithm for extracting presumed germline pathogenic variants in liquid genomic profiling: Insights from Database of Center for Cancer Genomics and Advanced Therapeutics (C-CAT)

**DOI:** 10.1101/2025.05.29.25327892

**Authors:** Satoyo Oda, Manami Matsukawa, Chikako Tomozawa, Noriko Tanabe, Tomoko Watanabe, Takafumi Koyama, Teruhiko Yoshida, Makoto Hirata

**Author notes:** Corresponding author: Makoto Hirata, Department of Genetic Medicine and Services, National Cancer Center Hospital 5-1-1 Tsukiji, Chuo-ku, Tokyo, 104-0045 Japa.

## Abstract

Liquid biopsy comprehensive genomic profiling (LB-CGP) testing is performed on circulating tumor DNA to detect tumor recurrence, predict prognosis, and select therapeutic agents. Pathogenic variants of germline origin in genes associated with hereditary tumor syndrome (HTS) can be simultaneously detected by CGP testing. Nonetheless, it is often challenging to differentiate whether the variants are of somatic or germline origin. The differentiation criteria were primarily based on the variant allele frequencies (VAFs). However, more evidence is needed to establish clear criteria, and it is often difficult to differentiate between variants based on VAF alone. In this study, using the national database of the Center for Cancer Genomics and Advanced Therapeutics, which accumulates real-world data on CGP testing in Japan, we analyzed 169,370 variants detected in 11,399 patients registered with FoundationOne Liquid CDx testing. By extracting the predominantly presumed somatic and germline variants, we established a criterion for VAF that could achieve high specificity and sensitivity. Further investigation into the detection status of other variants led to the development of an algorithm for differentiating somatic/germline variants of genes associated with HTSs. Based on this algorithm, 726 variants were extracted as presumed germline pathogenic variants among the 26 genes with high germline conversion rates in 710 patients in the study. This algorithm should help to discriminate with high accuracy whether the variants detected in LB-CGP tests are of somatic or germline origin, although further analyses are required to confirm the validity of this algorithm.

**Highlights:**

– The highly accurate VAF criterion for differentiating somatic and germline variants was determined using real-world data from more than 11,000 patients who underwent liquid biopsy CGP testing.
– To improve the specificity of the PGPV extraction, an additional criterion was defined: checking the status of other genomic alterations detected.
– Criteria for considering information other than VAFs associated with the variant under PGPV consideration were developed to improve the sensitivity of PGPV extraction.
– By integrating these criteria and previous evidence on germline conversion rates, a GeMSort algorithm was established.
– Based on this algorithm, 726 variants from 710 patients were extracted as PGPVs among 26 hereditary tumor syndrome-associated genes with high germline conversion rates.

## 1. Introduction

Liquid biopsy comprehensive genomic profiling (LB-CGP) testing, such as FoundationOne Liquid CDx and Guardant360 CDx, is currently reimbursed by the national health insurance system in Japan. The genomic findings from these tests are collected as real-world data by the Center for Cancer Genomics and Advanced Therapeutics (C-CAT)[1].

LB-CGP testing, which aims to identify somatic mutations in circulating tumor DNA (ctDNA), may also detect germline pathogenic variants as secondary findings, as these cannot be readily distinguished from somatic mutations[2–10]. Such potential germline findings from CGP testing may play a critical role in assessing hereditary cancer risk and guiding risk management strategies for patients and their relatives. Accordingly, presumed germline pathogenic variants (PGPVs) should be highlighted among the mutations listed in CGP test reports[4]. Previous studies investigating PGPVs in LB-CGP testing have shown that their extraction algorithms rely primarily on variant allele frequency (VAF)-based criteria[2, 5, 7–9, 11]. These studies contributed to the development of the LB-CGP operational guidelines for germline confirmatory testing of secondary findings in comprehensive tumor genomic profiling using circulating tumor DNA in the blood[12].

However, the application of these guidelines has shown limitations: some germline variants have been identified without meeting the criteria, while some somatic mutations have been incorrectly presumed to be germline due to the guidelines’ limited specificity. Currently, there is limited evidence to support more accurate algorithms for differentiating PGPVs.

In this study, we developed an algorithm for extracting PGPVs from LB-CGP testing, based on the LB-CGP operational guidelines and real-world databases collected by C-CAT.

## 2. Methods

### 2.1. Subjects and ethical approval

We extracted the variant data and clinical information of 11,407 patients who underwent FoundationOne Liquid CDx in Japan from the C-CAT repository databases ver. 20240621 on July 12, 2024[1]. For this research, the patient identification numbers were assigned an anonymized number as research case IDs. Approval was obtained from the institutional review board for the use of the C-CAT repository databases (#2019-301). The raw data for this study, including variants and clinical information, were obtained from the C-CAT Research Use Portal (https://www.ncc.go.jp/en/c_cat/use/index.html). Of the 11,407, two and three were excluded because of discrepancies in sex (prostate cancer in a female and uterine endometrial cancer in a male) and in CGP testing (variant data were registered from other CGP testing). In addition, three patients were excluded because the CGP testing reported no variants. Finally, 11,399 patients were included in the study (**Fig. 1A**).

**Fig. 1.**
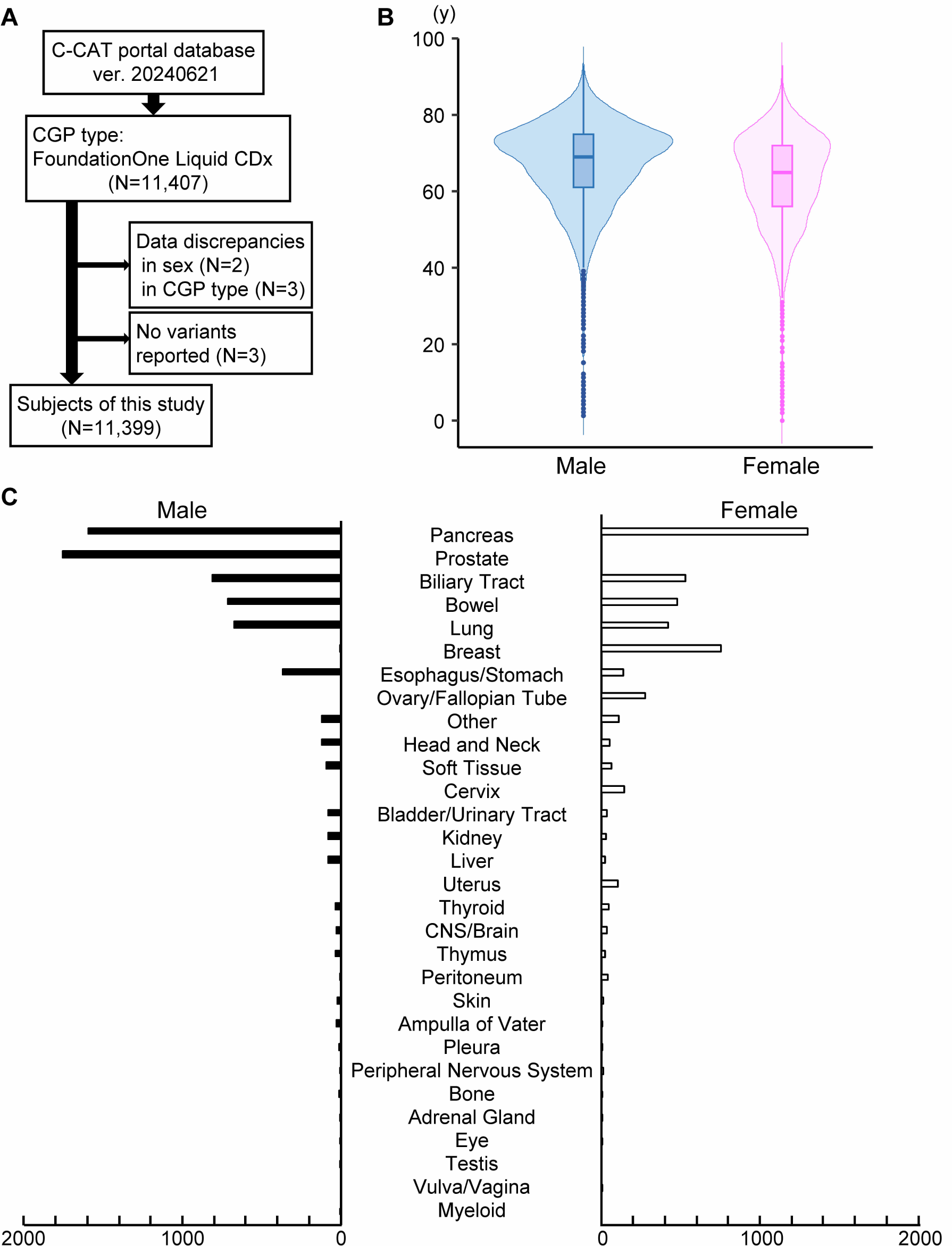
Clinical backgrounds of the subjects in this study. (**A**) Flowchart diagram of subjects’ selection. (**B**) Sex and age distribution of the subjects. The boxes represent the interquartile range, bounded by the first and third quartiles. The lines within the boxes denote the median age. (**C**) Frequency of cancer types in this study. Cancer types were classified according to OncoTree.

### 2.2. Variant evaluation

FoundationOne Liquid CDx reported 169,370 variants, including VUS, in 11,399 patients. We classified the variants into 11 categories: missense (132,310 variants), frameshift indel (14,602), stop gain (9,580), in-frame indel (6,428), splice site (5,260), promoter (575), stop loss (271), start loss (206), synonymous (77), intron (38) and exon deletion (23) (**Fig. S1A**). Variants of the *TP53* gene were the most frequently reported, followed by *DNMT3A*, *ATM*, *CHEK2*, and *APC* (**Fig. S1B**).

We re-evaluated variant pathogenicity according to the ACMG/AMP guidelines[13]. Briefly, we employed variant allele frequency from population databases, including Exome Aggregation Consortium (ExAC), the Genome Aggregation Database (gnomAD), and the Tohoku Medical Megabank Organization (ToMMo)[14–16], as well as case-control studies, previous experimental functional analyses, in-silico prediction tools such as SIFT, Polyphen2, LRT, MutationTaster2, and AlphaMissense[17–26], and variant databases, such as ClinVar[27] and COSMIC[28]. These resources were used for annotation to determine pathogenic and benign criteria and to assess the variant pathogenicity comprehensively.

In addition, for a comprehensive evaluation of the variants under certain conditions, quantile plots of VAF were constructed by plotting the VAF values of all variants of interest in ascending order, with all variants represented as one on the X-axis (**Fig. S1C**). VAF was categorized from C1 to C6 in increments of 0.2 (**Table S1**).

### 2.3. Selection and stratification of genes associated with hereditary tumor syndromes or oncogenic driver mutations

We selected 57 genes associated with hereditary tumor syndromes (HTSs) according to the list of secondary findings to be disclosed to patients according to the level of recommendation (ver. 4.2_20231003)[29]. Among the 57 genes, 26 were classified as having a high (>50%) germline conversion rate (GCR), four as moderate (10–50%), nine as low (<5%), and 11 as uncertain GCR. The germline conversion rate was based on previous evidence of the variant detection frequency in matched-pair CGP testing[30, 31]. The other genes, including *APC* and *TP53*, show low GCR in general but high or moderate GCR under specific conditions.

To analyze representative predominantly presumed germline variants (rPPGVs), we selected common polymorphisms identified with > 1% allele frequency in the Japanese population database (**Table S2**). In addition, to analyze representative predominantly presumed somatic variants (rPPSVs), we selected oncogenic hotspot mutations in *KRAS*, *PIK3CA*, *TERT*, *BRAF*, *NRAS*, and *EGFR*, which are frequently registered in the COSMIC database (**Fig. S2**).

### 2.4. Statistical Analysis

We used the R Commander to draw histograms, receiver operating characteristic (ROC) curves, and violin plots. Fisher’s exact test was used for comparative analysis of variant detection frequency between this study and the population database, using the R Commander. Continuous variables that did not follow a normal distribution were compared between the two independent groups using the Mann–Whitney U test. Fisher’s exact test for analysis of the carrier frequency of SNPs used a genome-wide significance level of 5 × 10^-8^ as the significance standard. For other statistical analyses, statistical significance was set at *P* < 0.05.

## 3. Results

### 3.1. Clinical backgrounds of the subjects

Of the 11,399 patients included in this study (**Fig. 1A**), patients in their 70s were the most frequent in both males and females (**Fig. 1B**), and those with pancreatic cancer were the most prevalent, followed by those with prostate, biliary duct, colorectal, and lung cancer (**Fig. 1C**), suggesting that this test is preferred for these cancer types. A total of 169,370 variants, including their allele frequencies, genome positions, and altered sequences, were reported in these patients and were categorized into six groups according to their allele frequencies, C1 to 6 (**Fig. S1C**). Variants with an allele frequency of 0.001 or more in population databases are rare (0.6%) in C1 variants but most frequent in C3 (52.8%) and C6 (48.8%). The variants on the X chromosome were more frequently reported in C5 (17.2%) or C6 (67.7%). These distributions supported the inference of the origin or status of variants according to the category (**Table S1**).

### 3.2. Threshold of variant allele frequency (VAF) for discriminating presumed germline variants

To determine the VAF threshold for discriminating presumed germline variants from somatic variants, we first extracted predominantly presumed somatic and germline variants among the 169,370 variants. We selected oncogenic hotspot mutations in *KRAS*, *PIK3CA*, *TERT*, *BRAF*, *NRAS*, and *EGFR*, which are frequently registered in the COSMIC database but not found in the population databases (**Fig. S2**) as rPPSVs. In addition, we extracted common polymorphisms identified with > 1% allele frequency in the Japanese population database as rPPGVs. The detection frequency of rPPGVs did not significantly differ from that of the population database at the genome-wide significance level (**Table S2**). The VAFs of rPPGVs accumulated around 0.5 (**Fig. 2A and Fig. S3**). These results indicated that the majority of rPPGVs were of germline origin. Using these variant data, we compared the distribution of the VAFs of rPPSVs and rPPGVs to gain insight into the threshold for discriminating presumed germline variants in LB-CGP testing. The VAFs of rPPSVs were relatively low, peaking below 0.05 and decaying rapidly, while those of rPPGVs were primarily detected in the variant category C3 (**Fig. 2A and Figs. S3-S4**). The sensitivity, specificity, and ROC curves indicated that 0.372 VAF had the highest area under the ROC curve (AUC, 0.974), with 0.978 sensitivity and 0.951 specificity (**Fig. 2B-C**). According to the previous studies or LB-CGP operational guidelines, 0.3 or more of VAF is the criterion for suspecting the variants as germline origin[2, 9, 12]. When the VAF criterion was set to 0.3, the sensitivity and specificity were 0.988 and 0.926, respectively.

**Fig. 2.**
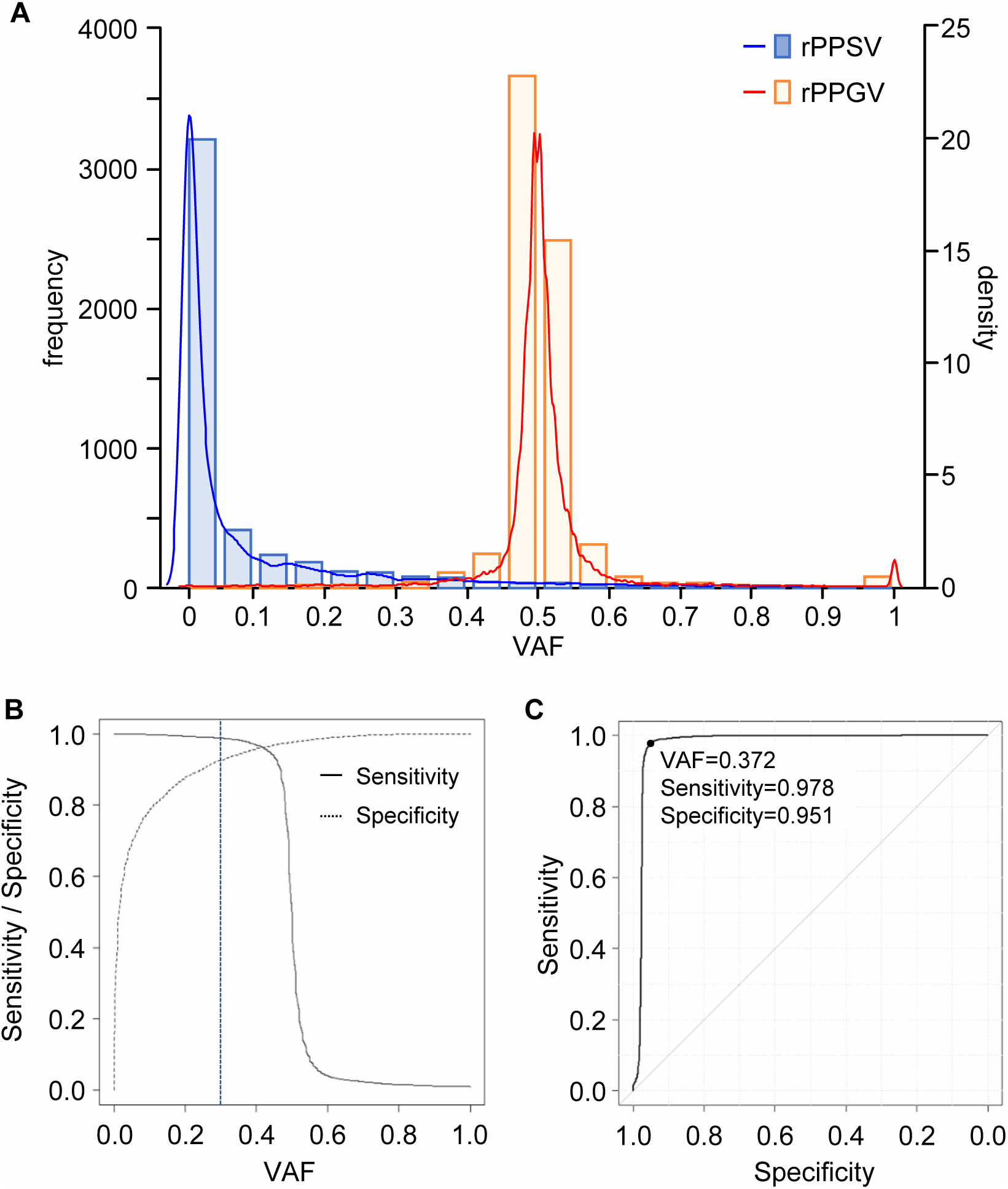
Threshold of variant allele frequency (VAF) to presume germline origin variants in liquid biopsy CGP testing. (**A**) Histograms and probability density distributions of VAF of representative predominantly presumed somatic variants (rPPSVs) and predominantly presumed germline variants (rPPGVs). Representative PPSVs were selected from frequent hotspot variants in oncogenes (**Fig. S2**). Representative PPGVs were selected from common SNPs, which were identified in more than 1% of the population database (**Table S2 - Fig. S3**). (**B, C**) Sensitivity/specificity (**B**) and receiver operating characteristic (ROC) (**C**) curves of VAF to predict germline origin variants, based on rPPSV and rPPGV VAF distribution. The area under the ROC curve was 0.974 (95%CI: 0.970-0.978).

### 3.3. Exclusion criteria of somatic mutations in the cases with increased somatic fraction (ISF)

Among 4,609 rPPSVs, 341 (7.4%) from 325 cases showed VAFs of 0.3 or more (high-VAF rPPSVs) (**Fig. S4**), and other variants that were strongly presumed to be of somatic origin were also observed to exhibit high VAFs. Accordingly, we defined cases with an increased somatic fraction (ISF) as those harboring somatic variants with elevated VAFs, and we sought to establish the criteria for the identification of such cases with the aim of excluding somatic variants with high VAFs.

Variants located on the X chromosome were excluded from criterion development due to their inconsistent VAF distributions compared to variants on autosomal chromosomes (**Table S1**). The variants with an allele frequency of 0.001 or more in population databases were also excluded, because most of the variants were suspected to be VUS or of germline origin. Thereafter, the remaining variants to be analyzed were defined as rare autosomal variants.

Cases harboring rPPSVs with high VAFs were classified as ISF. Comparative analysis between cases with and without high-VAF rPPSVs revealed that the cases with high-VAF rPPSVs more frequently carried rare autosomal variants categorized as C2, C4, and C5 in addition to high-VAF rPPSVs themselves. In contrast, the majority of the cases without high-VAF rPPSVs harbored fewer than those in such categories (**Fig. 3A, Fig. S5A, and Table S3**). No notable differences were observed for variants in C3 and C6, which were presumed to represent rare autosomal germline variants. These findings suggest that the presence of a C2, C4, or C5 rare autosomal variant may serve as a potent indicator of ISF.

**Fig. 3.**
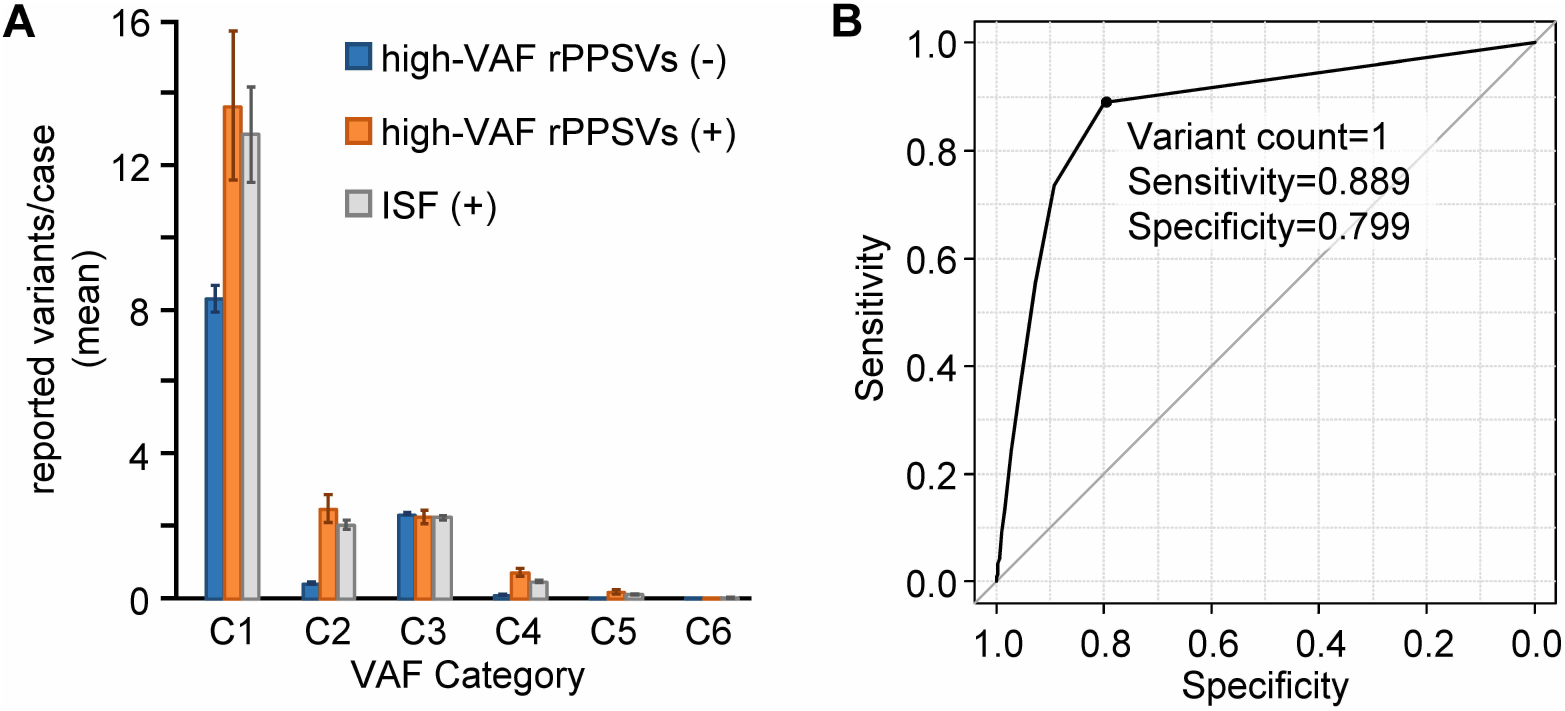
Characterization of increased somatic fraction (ISF). (**A**) Mean frequency of the rare autosomal variants per case with or without high-VAF rPPSVs by VAF categories. VAF was categorized from C1 to C6 in increments of 0.2 (**Table S1**). Error bars indicate 95% confidence interval. (**B**) Receiver-operating characteristic (ROC) curve of the variant count of VAF categories C2, 4, and 5 to discriminate the cases with high-VAF rPPSVs. The area under the ROC curve (AUC) of each category was 0.874 (95%CI 0.854-0.893).

To evaluate the discriminatory performance of these variant categories for ISF classification, we conducted ROC curve analysis using rPPSV-positive and rPPSV-negative cases. The combined count of C2, C4, and C5 variants yielded the highest AUC (0.874; 95%CI: 0.855-0.894), indicating robust discriminatory capability (**Fig. 3B-Fig. S5B, Table S4**). The optimal cut-off value, determined via the Youden index (variant count =1), achieved a sensitivity of 0.899 and specificity of 0.799 (**Fig. 3B**).

Based on these results, ISF cases were finally defined as those in which either a C2, C4, or C5 rare autosomal variant or a high-VAF rPPSV was detected. When comparing the high-VAF rPPSV and ISF cases, the detected variant counts by VAF category were almost comparable (**Fig.3A, Table S3**). According to this ISF definition, pathogenic variants in the HTS-associated genes with VAF ≥0.3, if detected with other C2, C4, or C5 rare autosomal variant(s) or high-VAF rPPSV(s), were excluded from further germline consideration, because they were more likely of somatic origin. Notably, because genes with low or uncertain GCR were presumed to have a low prior probability of harboring germline pathogenic variants, the additional filtering criterion was applied exclusively to variants in these genes, thereby improving the specificity of PGPV detection.

In 18 genes classified as low or uncertain germline conversion rate (GCR) genes by the LB-CGP operational guidelines, 52 pathogenic variants from 49 cases with ISF were detected at VAFs of 0.3 or higher by FoundationOne Liquid CDx. All the cases met the ISF criteria; none had a phenotype or family history that strongly suggested an associated hereditary tumor syndrome (**Table S5**).

### 3.4. Additional identification of presumed germline variants with lower VAF

Because variants with longer insertions or deletions (indels) tended to show lower VAF due to mapping errors, common polymorphisms with long indels (more than 10 bp), *RAD54L* C391fs, *KMT2D* S696_D704del, *NOTCH3* R10_R13del, *GNAS* P453_D464del, *TSC2* D1690fs, and *MTOR* T1830_1833del were selected (**Fig. S6A and Table S2**) to examine their VAF distributions. The VAF distributions increased the C2 proportion compared to other common SNPs or short indel polymorphisms (**Figs. S3, S6B**). Because 98.9% of the variants had VAFs of 0.2 or higher, a VAF threshold of 0.2 is defined for identifying presumed germline variants for such long indel variants.

Somatic reversion mutations that arise in cases with *BRCA1* or *BRCA2* (*BRCA1/2*) germline pathogenic variants are one of the mechanisms of drug resistance through prolonged administration of platinum-based drugs or Poly(ADP-ribose) polymerase (PARP) inhibitors[10, 32, 33]. Such somatic mutations can also decrease the allele frequency of *BRCA1/2* pathogenic germline variants (**Table S6**, case# 5198). Hence, another node to verify whether such presumed somatic reversion mutations in *BRCA1/2* were detected was added to the algorithm.

Pathogenic variants with VAFs between 0.2 and 0.3 in high-GCR genes were extracted to examine if they were of germline origin. No cases with such variants, except for those with other PGPVs with VAFs of 0.3 or higher, were suspected of HTSs. Most of the cases met the ISF criteria (**Table S6**).

### 3.5. Developing a germline and somatic mutation sorting (GeMSort) algorithm for extracting presumed germline pathogenic variants

Finally, an algorithm for extracting PGPVs from LB-CGP testing (GeMSort) was formulated by integrating these results with the LB-CGP operational guidelines (**Fig. 4**).

**Fig. 4.**
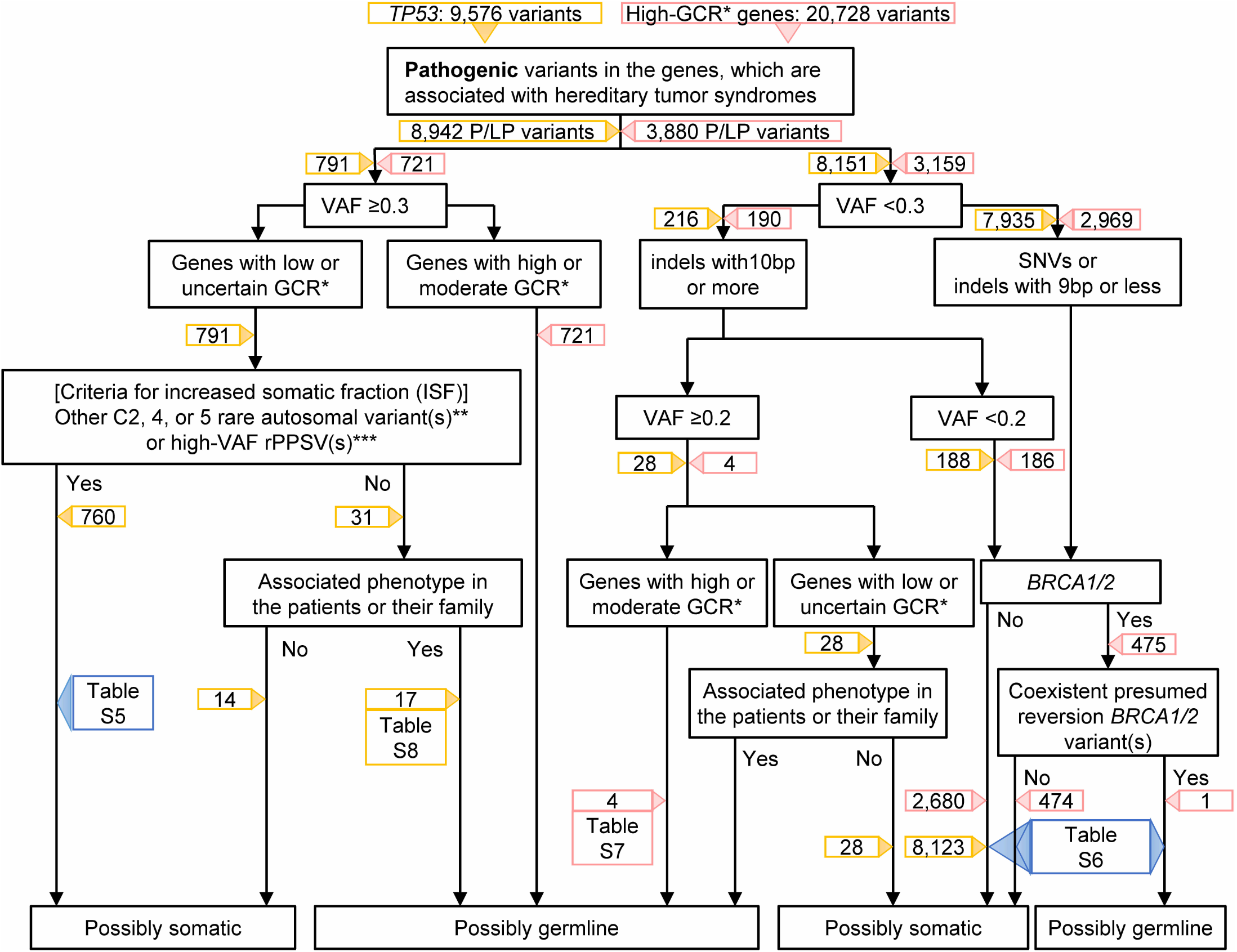
Germline and Somatic Mutation Sorting (GeMSort) Algorithm for extracting PGPVs in LB-CGP testing. Yellow and light red boxes indicate the number of variants, sorted by each node, in TP53 and high-GCR genes, respectively. * GCR, Germline Conversion Rate, ** Variant categories, defined in **Table S1**, *** representative predominantly presumed somatic variants, described in **Fig. S2**.

Using the GeMSort algorithm, variants of representative genes associated with HTS were screened to discriminate between somatic and germline variants. Of the 26 high-GCR genes listed in the LB-CGP operational guidelines, 20,728 variants from all the subjects were reported using FoundationOne Liquid CDx. A total of 3,880 variants were classified as pathogenic or likely pathogenic. Among the pathogenic or likely pathogenic variants, 721 with 0.3 or more VAF were incorporated into PGPVs, while five variants of 3,159 with less than 0.3 VAF were considered as PGPVs (**Figs. 4-5A-Tables S6-S7**). The variants in *BRCA2* were the most frequently identified PGPVs, followed by *ATM* and *BRCA1* variants (**Fig. 5B**). Regarding cancer type, PGPVs were detected in more than 20% of the patients of peritoneal and ovarian/fallopian tube cancer and in more than 10% of patients of adrenal and breast cancer (**Fig. 5C**). In the *TP53* gene, which generally shows a low GCR except under specific conditions, 9,576 variants have been reported. Overall, 8,942 variants were classified as pathogenic or likely pathogenic. All 8,151 variants with VAF < 30% were judged to be possibly somatic. In addition, among the 791 with a VAF of 30% or higher, 735 variants accompanied by ISF were classified as possibly somatic. In contrast, 17 of the remaining 56 variants were classified as possibly germline because of the phenotype associated with the pathogenic variants (**Fig. 4-Table S8**).

**Fig. 5.**
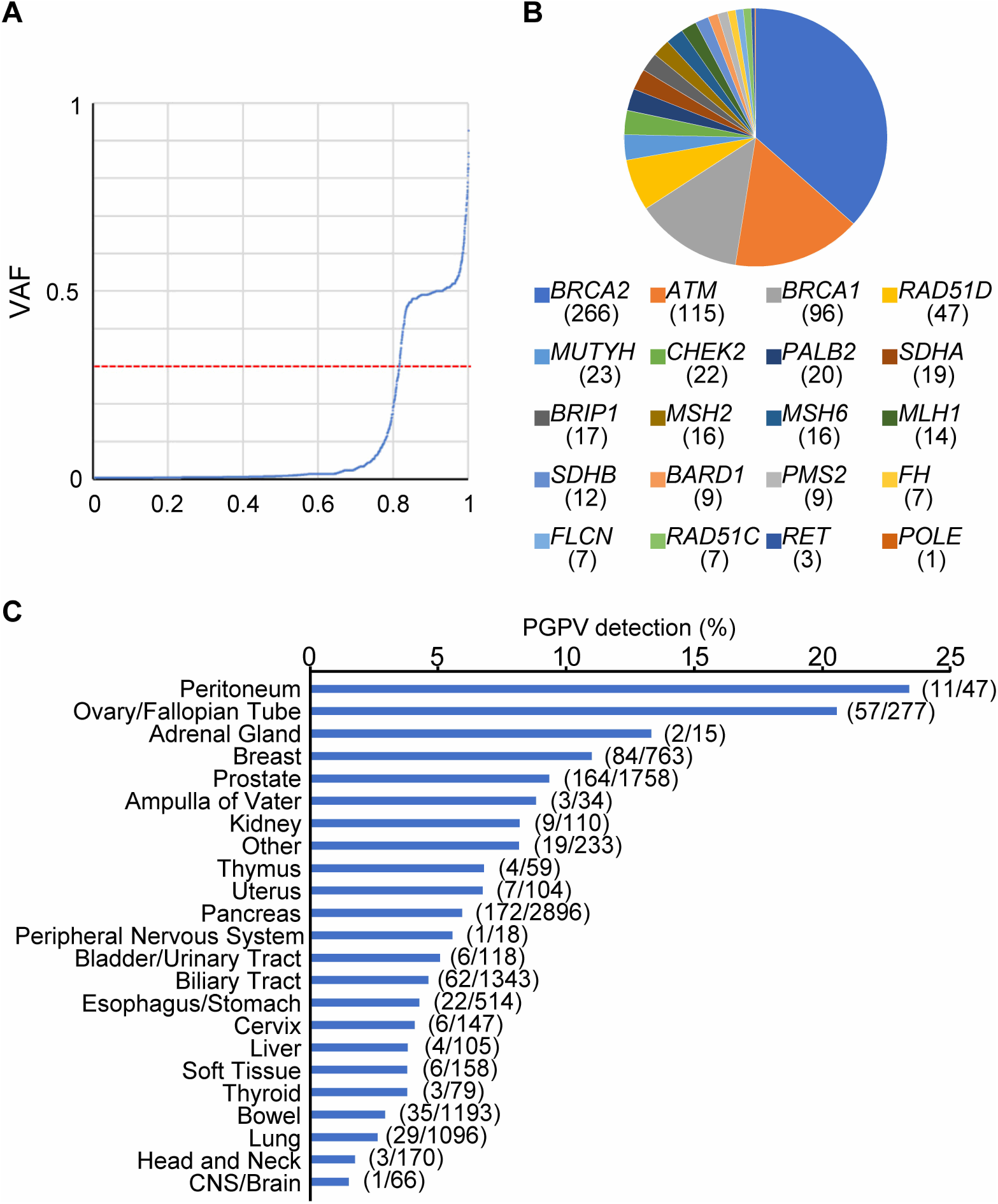
Presumed germline pathogenic variants of high-GCR genes extracted by GeMSort algorithm. (**A**) A quantile plot of VAF for pathogenic/likely pathogenic variants in high germline-conversion-rate (GCR) genes. This curve is constructed by plotting the VAF of 3,880 variants in ascending order, with 3,880 variants represented as 1 on the X-axis. High GCR genes were defined elsewhere[12, 29]. (**B**) Frequency of PGPVs of high-GCR genes. Overall, 726 variants were identified in 710 patients according to the GeMSort algorithm. (**C**) Frequency of patients with PGPVs in cancer types. The fraction in parentheses indicates the patients with PGPV(s) / subject cases.

## 4. Discussion

This study refined the LB-CGP operational guidelines for selecting PGPVs using C-CAT national databases and established a novel germline and somatic mutation sorting (GeMSort) algorithm (**Fig. 4**) for extracting PGPVs in LB-CGP testing.

Several studies have examined germline-origin variants using LB-CGP testing[2, 5, 7–9, 11]. In some research, presumed germline variants were defined as those in which the VAF was between 0.4 and 0.6[7, 8], while others discriminate the presumed germline variants with a threshold of at least 0.3 of VAF[2, 9]. This study demonstrated that a VAF threshold of 0.372 yielded the highest AUC for distinguishing between presumed germline and somatic variants by analyzing the rPPGVs and rPPSVs (**Fig. 2**). However, a lower VAF threshold of 0.3, which improves sensitivity, seems more suitable for clinical application because of the increasing relevance of identifying actionable germline findings and the critical need to ensure that these findings are not missed.

Conversely, a VAF threshold of 0.3 resulted in reduced specificity: approximately 7% of rPPSVs were detected with a VAF of 0.3 or higher (**Fig. 2**). Elevated VAFs typically correlated with an increased somatic fraction, ISF. Certain patients with ISF exhibited two or more pathogenic variants with higher VAF in cancer predisposition genes with low or uncertain GCR without any apparent phenotype or family history of HTS, suggesting that these variants were somatic mutations (**Table S5**). Notably, for genes with a low GCR, where the likelihood of somatic origin is presumed high, utilizing ISF criteria to refine the identification of PGPVs in these genes could be advantageous.

FoundationOne Liquid CDx often detects an elevated tumor fraction. The tumor fraction estimates the percentage of circulating tumor DNA based on the observed level of aneuploid instability, using the frequencies of approximately 1,000 SNP sites across the genome[34]. Therefore, this parameter can also estimate the ISF status to some extent. However, in some cases, the tumor fraction could not be determined because of the absence of aneuploidy. In addition, it cannot evaluate variants associated with clonal hematopoiesis (CH). Some CH genes, including *ATM, CHEK2*, and *TP53*, are associated with HTS[35–38]. *TP53* pathogenic variants with 0.3 or more of VAF are sometimes detected in aged patients without an apparent family history of cancers. Even in such cases, the ISF criterion can be a potent indicator for discriminating PGPVs from CH-derived variants. Hence, the ISF criterion may be clinically useful for excluding somatic (tumor- and CH-derived) mutations.

Reversion mutations have been identified in patients with *BRCA1/2* pathogenic variants where prolonged treatment with platinum-based chemotherapy or PARP inhibitors[10, 32, 33]. PARP inhibitors may also be used in patients with homologous recombination (HR) deficiencies associated with other germline pathogenic variants of HR-associated genes, such as *ATM*, *BRIP1*, *BARD1*, *PALB2*, and CHEK2[39]. Notably, the other HR-associated genes were not included in the node with reversion mutations in the GeMSort algorithm. A previous study identified a *RAD51D* secondary resistance mutation induced by exposure to a PARP inhibitor in a patient with a germline *RAD51D* pathogenic variant[40]. The future prevalence of PARP inhibitors in patients with HRD may also contribute to the increased frequency of reversion mutations in other HR-associated genes. Further accumulation of evidence regarding reversion mutations is essential for revising the reversion mutation node in the algorithm.

The limitation of the current study is the absence of data to confirm the origin of the PGPVs using non-tumor tissue, as the national health insurance in Japan does not currently cover the validation of normal tissue testing, even when such presumed pathogenic variants are identified. The availability of cancer history data in C-CAT databases is also limited. Another limitation is the exclusive use of FoundationOne Liquid CDx in this study. The applicability of these algorithms to other LB-CGP testing needs further examination.

In conclusion, we established the GeMSort algorithm, which is versatile and useful, based on real-world data from over 10,000 patients. Future validation studies on the algorithm’s utility should include results from LB-CGP and confirmatory germline testing.

## Abbreviations

CGP: comprehensive genomic profiling
C-CAT: Center for Cancer Genomics and Advanced Therapeutics
PGPVs: presumed germline pathogenic variants
ExAC: Exome Aggregation Consortium
gnomAD: The Genome Aggregation Database (gnomAD)
ToMMo: Tohoku Medical Megabank Organization
VAF: variant allele frequency
GCR: germline conversion rate
rPPGVs: representative predominantly presumed germline variants
rPPSVs: representative predominantly presumed somatic variants
ROC: receiver operating characteristic
AUC: area under the ROC curve
ISF: increased somatic fraction
GeMSort algorithm: a germline and somatic mutation sorting algorithm
PARP: Poly(ADP-ribose) polymerase

## Data Availability

All data used in the present study are available online at the C-CAT Research Use Portal (https://www.ncc.go.jp/en/c_cat/use/index.html).
All data produced in the present study are contained in the manuscript.

## Acknowledgments

We express our gratitude to the patients and their families who participated in this study. We are grateful for the support of our colleagues involved with the molecular tumor board of CGP testing at the National Cancer Center Hospital.

## Author Contributions

S.O. and M.H. conceptualized, designed the study, and drafted the manuscript. S.O., M.M., C.T., N.T., and T.W. acquired, analyzed, and interpreted the data. M.M., C.T., N.T., T.W., T.K., and T.Y. critically revised the manuscript for important intellectual content. T.Y. and M.H. obtained funding. T.K., T.Y., and M.H. supervised the study.

## Funding

This work was supported by the National Cancer Center Research and Development Funds (2023-A-18) and the Health Labour Sciences Research Grant (23EA1037).

## Competing interests

The authors have no conflict of interest.

## Supplementary Figures

**Fig. S1.** Types and allele frequency distribution of the variants in this study. (**A**) Distribution of the variant types. The numbers in parentheses indicate the frequency of respective variant types. A total of 169,370 variants were included in this study. (**B**) Cumulative bar chart showing the number of the mutations by type in frequently reported genes. Overall, 32 genes with more than 1,000 reported variants were selected. (**C**) A quantile plot of variant allele frequency (VAF) of all variants included in this study. The VAF values of all variants were plotted in ascending order, with 169,370 variants represented as 1 on the X-axis. VAF was categorized from C1 to C6 in increments of 0.2 (**Table S1**).

**Fig. S2.** Details and frequency of representative predominantly presumed somatic variants (rPPSVs). Pie charts represent the distribution of hotspot mutations in the *KRAS*, *PIK3CA*, *TERT*, *BRAF*, *NRAS*, and *EGFR* genes. The numbers in parentheses indicate the detected variant numbers of respective gene hotspots.

**Fig. S3.** Quantile plots of VAF of representative predominantly presumed germline variants (rPPGVs). The VAF values of common polymorphisms of respective genes were plotted in ascending order, with all mutations represented as 1 on the X-axis. Common polymorphism variants (**Table S2**) were selected as rPPGVs. The numbers in parentheses indicate the numbers of respective gene variants. Red dotted lines on 0.3 of VAF indicate the criteria for extracting presumed germline variants.

**Fig. S4.** Quantile plots of VAFs of representative predominantly presumed somatic variants (rPPSVs). The VAF values of hotspot somatic mutations of respective genes were plotted in ascending order, with all mutations represented as 1 on the X-axis. *KRAS*, *PIK3CA*, *TERT*, *BRAF*, *NRAS*, and *EGFR* hotspot mutations (**Fig. S2**) were selected as rPPSVs. The numbers in parentheses indicate the numbers of respective gene variants. Red dotted lines on 0.3 of VAF indicate the criteria for predicting presumed germline variants. The *EGFR* T790M variant, which is detected at a 4.6×10^-5^ allele frequency in gnomAD v4, is included in this analysis because the variant is not identified in Japanese population databases.

**Fig. S5.** Development of the definition of increased somatic fraction (ISF). (**A**) A quantile plot of VAF of the cases with or without high-VAF rPPSVs. High-VAF was defined as a variant-allele frequency of more than 0.3. Overall, 6,259 variants from 325 cases with high-VAF PPSVs and 123,294 from 11,074 without high-VAF PPSVs were plotted, respectively. (**B**) Receiver-operating characteristic (ROC) curve of the variant count of VAF categories C2, 4, and 5 to discriminate the cases with high-VAF rPPSVs. The area under the ROC curve (AUC) of each category was described in **Table S4**.

**Fig. S6.** Relief extraction of presumed germline variants with decreased VAF. (**A**) Frequency of common polymorphisms with long indels. A total of six common polymorphisms were selected, each characterized by an insertion or deletion of 10bp or more. The parentheses indicate the number of cases and indel length of respective variants included in this study. (**B**) A quantile plot of VAF of common polymorphisms with long indels. VAF values of the six common polymorphisms with more than 10bp indels (**Table S2**) were plotted.

**Table S1.** Categorical classification and number of variants reported in this study.

**Table S2.** Predominantly presumed germline variants (PPGVs) detected in liquid CGP testing.

**Table S3.** Frequency of variants classified by VAF category in the cases with increased somatic fraction (ISF) or associated conditions.

**Table S4.** Criteria for variant detection numbers by category identified in each case to extract cases with high-VAF rPPSVs.

**Table S5.** Cases where pathogenic variants in genes with low or uncertain GCR were identified in association with ISF

**Table S6.** Cases with presumed somatic and germline pathogenic variants with 0.2≤VAF<0.3 in high GCR genes.

**Table S7.** Cases with presumed germline pathogenic variants with long indel and 0.2≤VAF<0.3.

**Table S8.** Cases with presumed germline pathogenic variants of *TP53*, selected by the GeMSort algorithm.

## Notes

### Competing Interest Statement

The authors have declared no competing interest.

### Author Declarations

IRB of National Cancer Center, Japan gave ethical approval for this work (#2019-301).

