## Supplemental_Figures_1-6 for "A germline and somatic mutation sorting (GeMSort) algorithm for extracting presumed germline pathogenic variants in liquid genomic profiling: Insights from Database of Center for Cancer Genomics and Advanced Therapeutics (C-CAT)"

### Figure S1

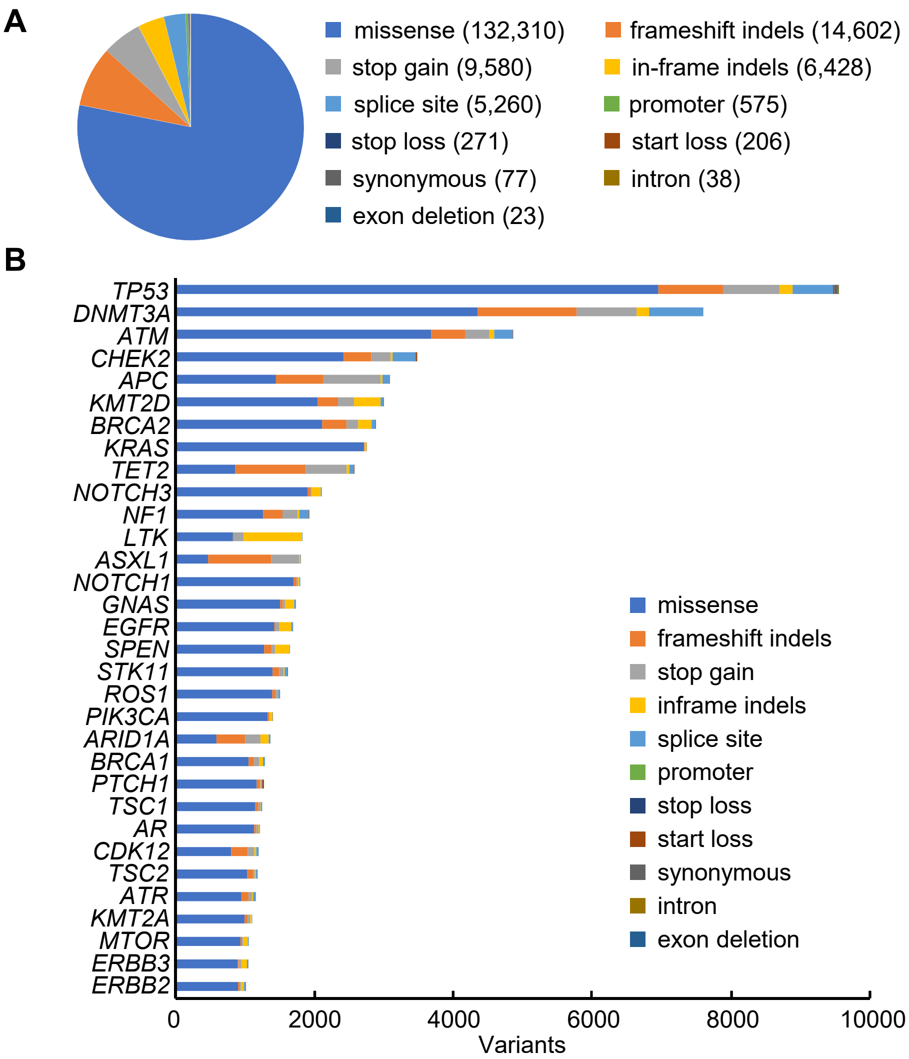


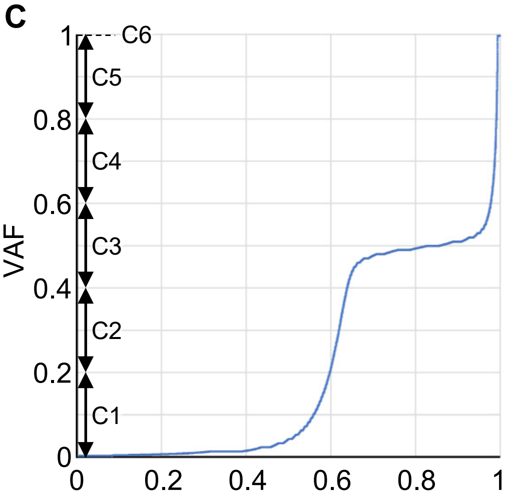


**Fig. S1. Types and allele frequency distribution of the variants in this study.** **(A)** Distribution of the variant types. The numbers in parentheses indicate the frequency of respective variant types. A total of 169,370 variants were included in this study. **(B)** Cumulative bar chart showing the number of the mutations by type in frequently reported genes. Overall, 32 genes with more than 1,000 reported variants were selected. **(C)** A quantile plot of variant allele frequency (VAF) of all variants included in this study. The VAF values of all variants were plotted in ascending order, with 169,370 variants represented as 1 on the X-axis. VAF was categorized from C1 to C6 in increments of 0.2 (**Table S1**).

**Figure S2**

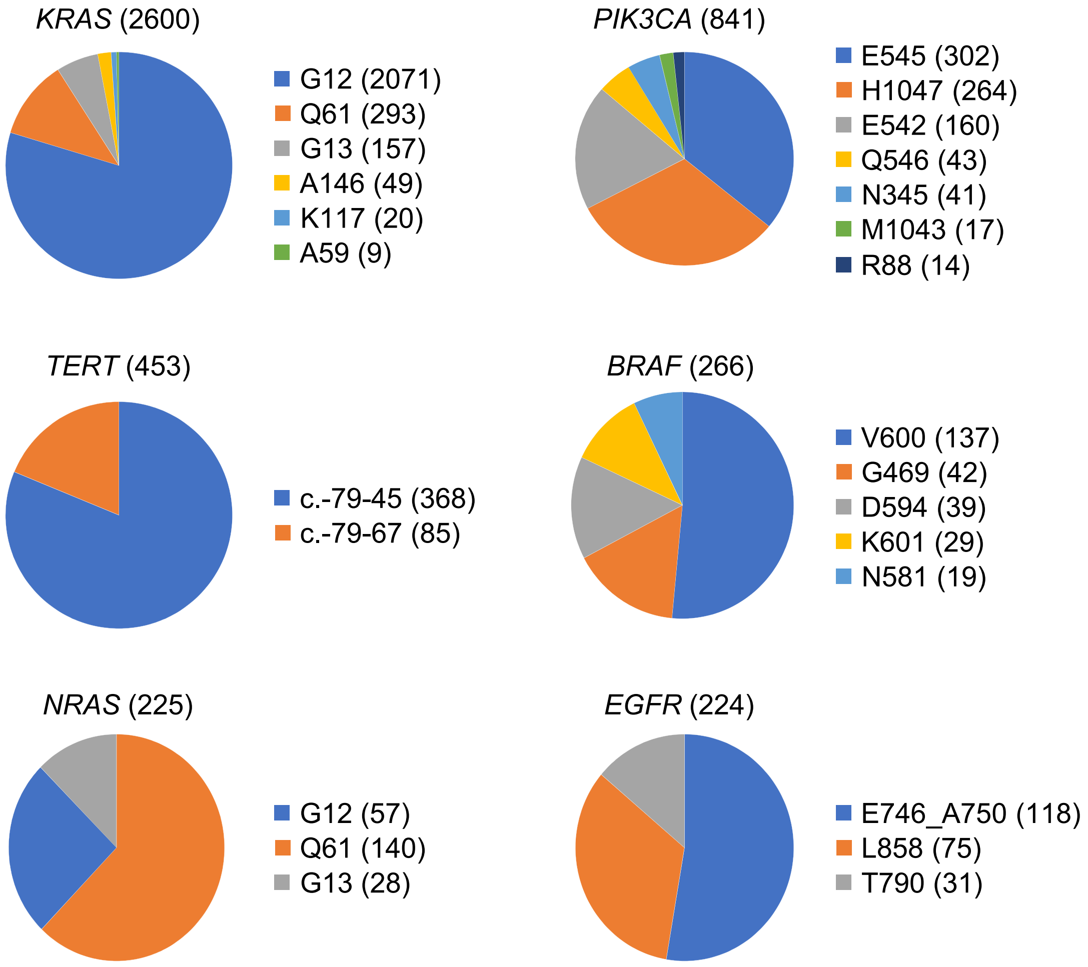


**Fig. S2. Details and frequency of representative predominantly presumed somatic variants (rPPSVs).** Pie charts represent the distribution of hotspot mutations in the *KRAS*, *PIK3CA*, *TERT*, *BRAF*, *NRAS*, and *EGFR* genes. The numbers in parentheses indicate the detected variant numbers of respective gene hotspots.

**Figure S3**

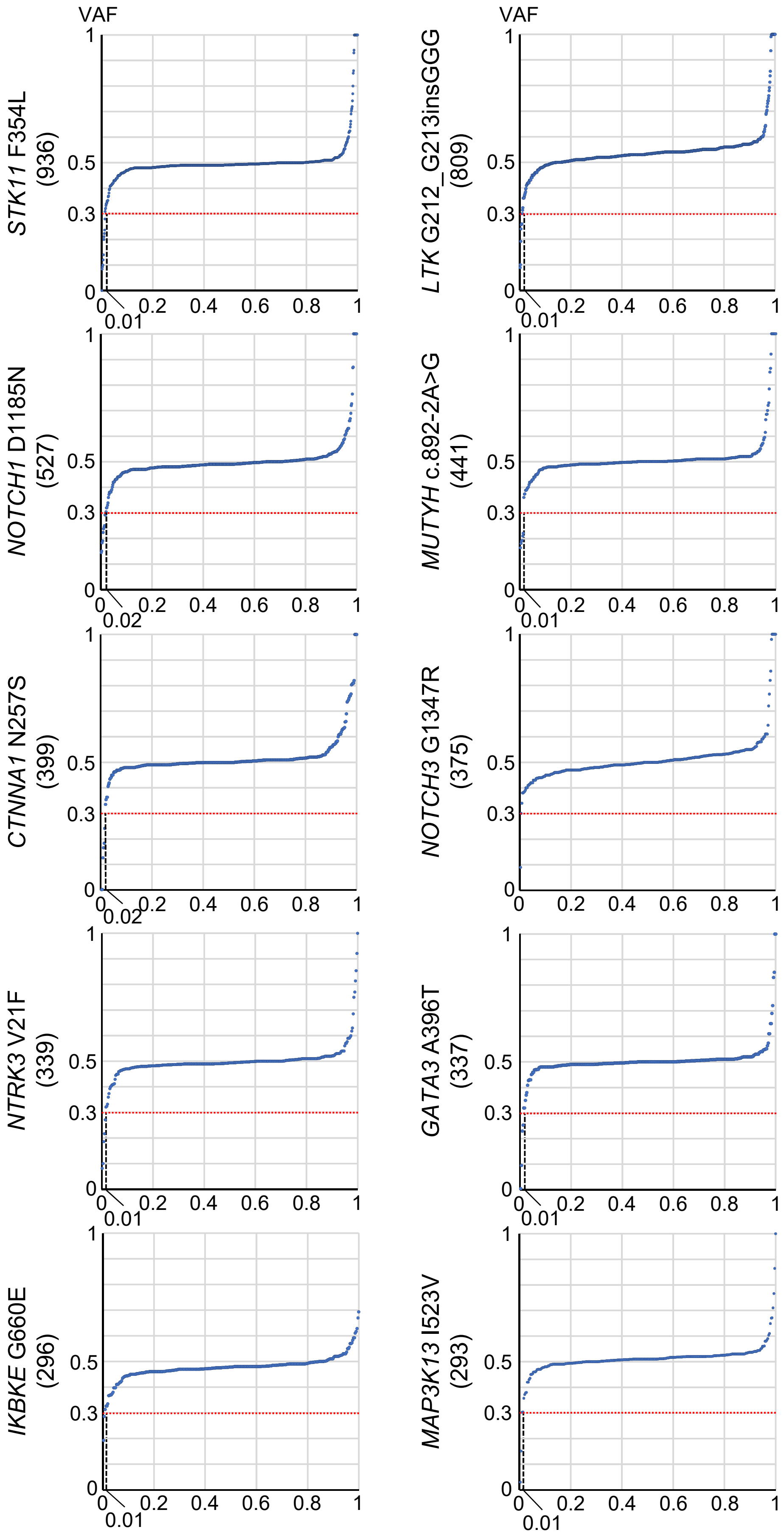

VAF
STK11 F354L (936)
LTK G212_G213insGGG (809)
NOTCH1 D1185N (527)
MUTYH c.892-2A>G (441)
CTNNA1 N257S (399)
NOTCH3 G1347R (375)
NTRK3 V21F (339)
GATA3 A396T (337)
IKBKE G660E (296)
MAP3K13 I523V (293)
0.01
0.02

**Figure S3_continued**

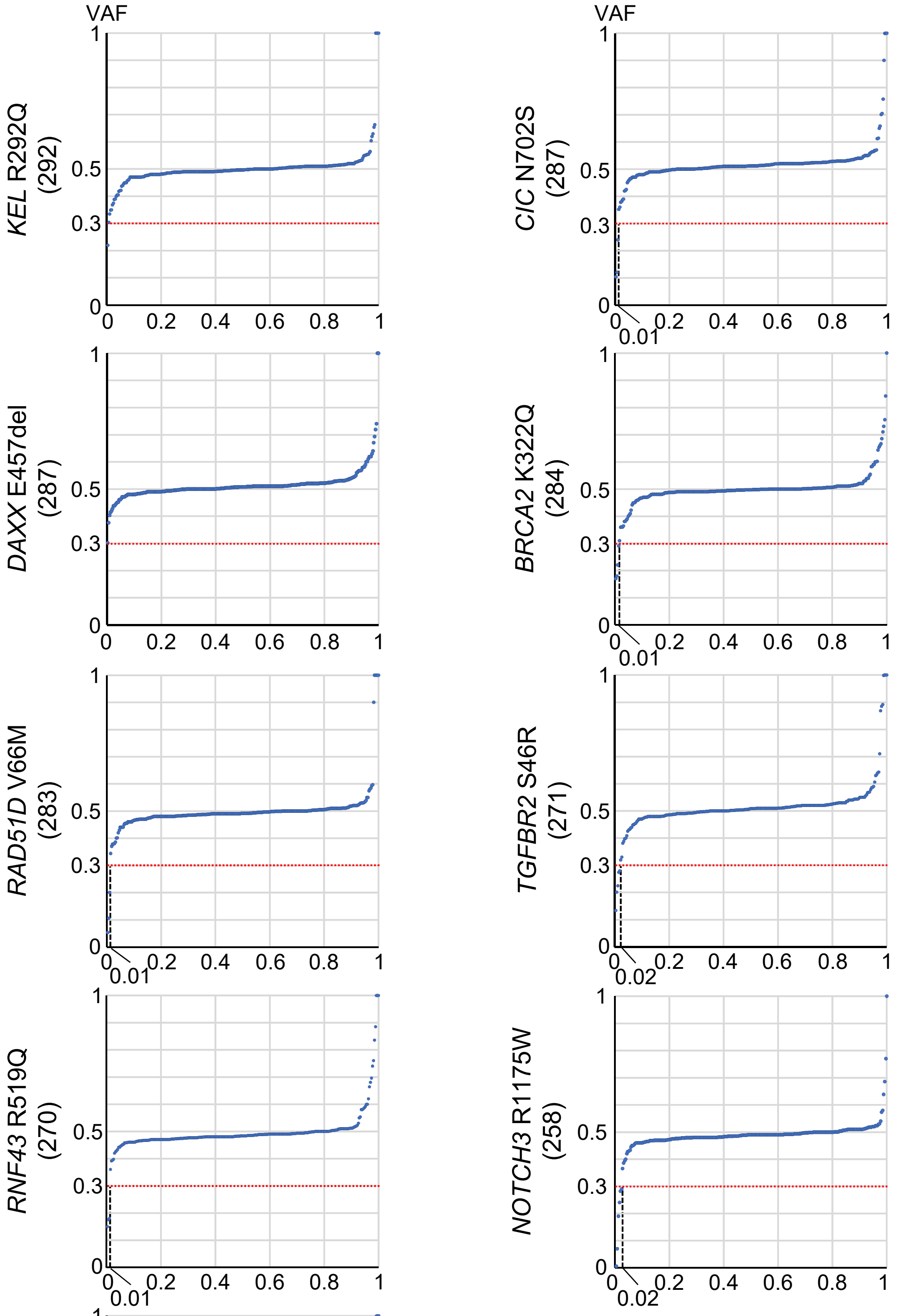


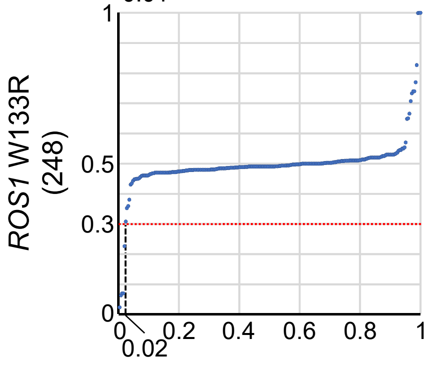


**Fig. S3. Quantile plots of VAF of representative predominantly presumed germline variants (rPPGVs).** The VAF values of common polymorphisms of respective genes were plotted in ascending order, with all mutations represented as 1 on the X-axis. Common polymorphism variants (**Table S2**) were selected as rPPGVs. The numbers in parentheses indicate the numbers of respective gene variants. Red dotted lines on 0.3 of VAF indicate the criteria for extracting presumed germline variants.

**Figure S4**

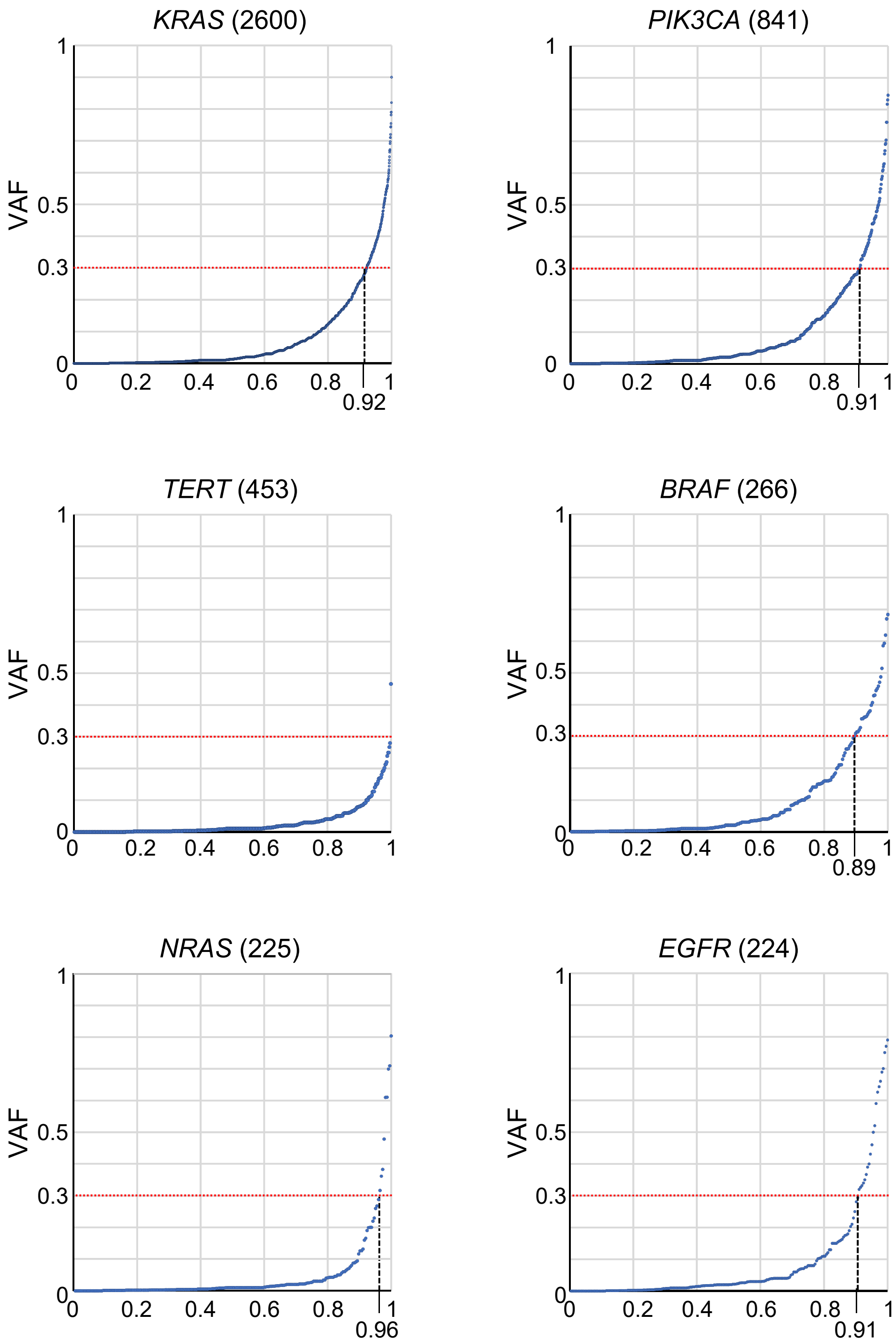


**Fig. S4. Quantile plots of VAFs of representative predominantly presumed somatic variants (rPPSVs).** The VAF values of hotspot somatic mutations of respective genes were plotted in ascending order, with all mutations represented as 1 on the X-axis. *KRAS*, *PIK3CA*, *TERT*, *BRAF*, *NRAS*, and *EGFR* hotspot mutations (**Fig. S2**) were selected as rPPSVs. The numbers in parentheses indicate the numbers of respective gene variants. Red dotted lines on 0.3 of VAF indicate the criteria for predicting presumed germline variants. The *EGFR* T790M variant, which is detected at a 4.6x10$^{-5}$ allele frequency in gnomAD v4, is included in this analysis because the variant is not identified in Japanese population databases.

#### Figure S5

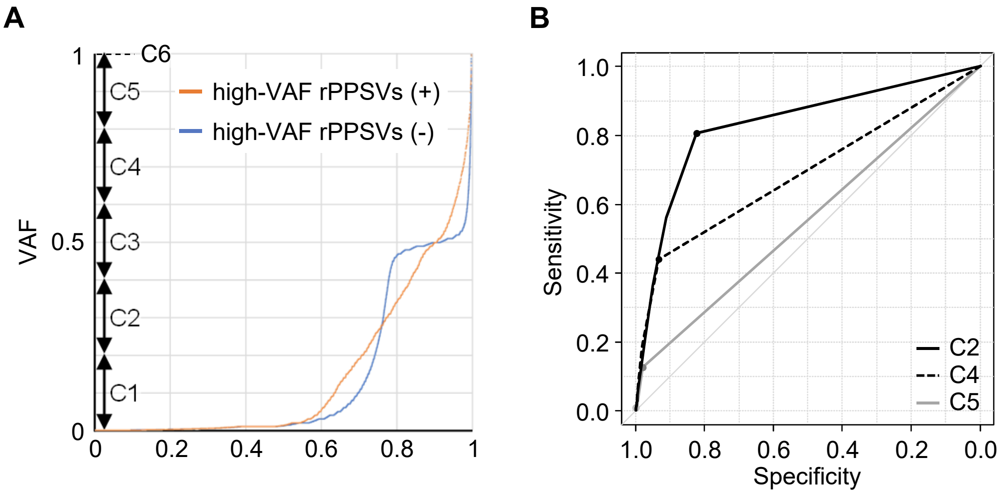


**Fig. S5. Development of the definition of increased somatic fraction (ISF).** **(A)** A quantile plot of VAF of the cases with or without high-VAF rPPSVs. High-VAF was defined as a variant-allele frequency of more than 0.3. Overall, 6,259 variants from 325 cases with high-VAF PPSVs and 123,294 from 11,074 without high-VAF PPSVs were plotted, respectively. **(B)** Receiver-operating characteristic (ROC) curve of the variant count of VAF categories C2, 4, and 5 to discriminate the cases with high-VAF rPPSVs. The area under the ROC curve (AUC) of each category was described in **Table S4**.

### Figure S6

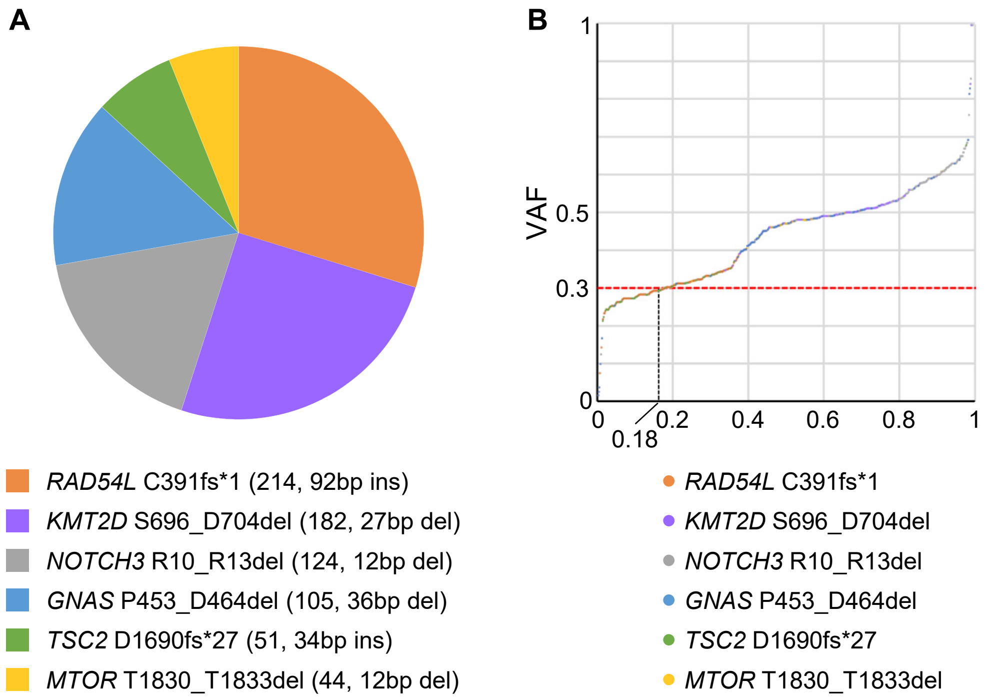


**Fig. S6. Relief extraction of presumed germline variants with decreased VAF.** **(A)** Frequency of common polymorphisms with long indels. A total of six common polymorphisms were selected, each characterized by an insertion or deletion of 10bp or more. The parentheses indicate the number of cases and indel length of respective variants included in this study. **(B)** A quantile plot of VAF of common polymorphisms with long indels. VAF values of the six common polymorphisms with more than 10bp indels (**Table S2**) were plotted.
