## Supplemental_Tables_1-8 for "A germline and somatic mutation sorting (GeMSort) algorithm for extracting presumed germline pathogenic variants in liquid genomic profiling: Insights from Database of Center for Cancer Genomics and Advanced Therapeutics (C-CAT)"

**Table S1**

**Table S1. Categorical classification and number of variants reported in this study**

| Category | Definition | Number of variants (rare autosomal variants*) | Number (%) of variants, with 0.001 or more of allele frequency in population databases** | Number (%) of variants on X chromosome | Major presumed status of the variants |
|---|---|---|---|---|---|
| C1 | 0≤VAF<0.2 | 100,976 (96,023) | 580 (0.6%) | 4,382 (4.3%) | Somatic variants |
| C2 | 0.2≤VAF<0.4 | 6,923 (5,471) | 1,182 (17.1%) | 278 (4.0%) | Somatic variants with increased somatic fraction, or Germline variants with decreased mapping reads |
| C3 | 0.4≤VAF<0.6 | 57,787 (26,724) | 30,503 (52.8%) | 859 (1.5%) | Heterozygous germline polymorphisms |
| C4 | 0.6≤VAF<0.8 | 2,307 (1,307) | 935 (40.5%) | 84 (8.9%) | Somatic or germline variants with loss of heterozygosity |
| C5 | 0.8≤VAF<1.0 | 565 (293) | 198 (35.0%) | 97 (17.2%) | Somatic or germline variants with loss of heterozygosity, germline/somatic variants on X chromosome, or homozygous germline polymorphisms with somatic mutation or sequencing error |
| C6 | AF=1.0 | 812 (76) | 396 (48.8%) | 550 (67.7%) | Homozygous germline polymorphisms or germline polymorphisms on X chromosome |

* Variants excluding those with 0.001 or more of allele frequency in population databases or located on X chromosome

** Three population databases are available in C-CAT databases; ToMMo, ExAC_eas and 1000 genome_eas

**Table S2. Predominantly presumed germline variants (PPGVs) detected in liquid CGP testing.**

| Variants | dbSNP ID | Carrier freq in this study | Carrier freq in ToMMo* | Allele freq in ToMMo* | Fisher's exact test | OR |
|---|---|---|---|---|---|---|
| *STK11* F354L | rs59912467 | 0.0821 | 0.0897 | 0.045921 | 4.66.E-04 | 0.91 |
| *LTK* G212_G213insGGG | rs756867166 | 0.0710 | 0.0622 | 0.031731 | 4.66.E-05 | 1.15 |
| *NOTCH1* D1185N | rs548083258 | 0.0462 | 0.0541 | 0.027412 | 4.50.E-05 | 0.85 |
| *MUTYH* c.892-2A>G | rs77542170 | 0.0387 | 0.0400 | 0.020229 | 1.73.E-02 | 0.97 |
| *CTNNA1* N257S | rs78627784 | 0.0350 | 0.0375 | 0.018931 | 9.45.E-03 | 0.93 |
| *NOTCH3* G1347R | rs1344432803 | 0.0329 | 0.0362 | 0.018371 | 5.10.E-03 | 0.91 |
| *NTRK3* V21F | rs200822610 | 0.0297 | 0.0274 | 0.013876 | 9.71.E-03 | 1.09 |
| *GATA3* A396T | rs200935603 | 0.0296 | 0.0321 | 0.016206 | 8.70.E-03 | 0.92 |
| *IKBKE* G660E | rs55822317 | 0.0260 | 0.0293 | 0.014806 | 3.89.E-03 | 0.88 |
| *MAP3K13* I523V | rs201038046 | 0.0257 | 0.0249 | 0.012596 | 2.32.E-02 | 1.03 |
| *KEL* R292Q | rs201698610 | 0.0256 | 0.0312 | 0.015708 | 1.30.E-04 | 0.82 |
| *CIC* N702S | rs202135824 | 0.0252 | 0.0291 | 0.014677 | 1.66.E-03 | 0.86 |
| *DAXX* E457del | rs200567881 | 0.0252 | 0.0242 | 0.012182 | 2.21.E-02 | 1.04 |
| *BRCA2* K322Q | rs11571640 | 0.0249 | 0.0240 | 0.01212 | 2.26.E-02 | 1.04 |
| *RAD51D* V66M | rs56026142 | 0.0248 | 0.0245 | 0.012256 | 2.59.E-02 | 1.01 |
| *TGFBR2* S46R | rs200111443 | 0.0238 | 0.0303 | 0.015331 | 1.55.E-05 | 0.78 |
| *RNF43* R519Q | rs200111443 | 0.0237 | 0.0306 | 0.015331 | 6.06.E-06 | 0.77 |
| *NOTCH3* R1175W | rs200504060 | 0.0226 | 0.0223 | 0.011178 | 2.69.E-02 | 1.02 |
| *ROS1* W133R | rs201807486 | 0.0218 | 0.0263 | 0.013324 | 4.60.E-04 | 0.82 |
| *RAD54L* C391fs*1 | rs1557706222 | 0.0188 | 0.0139 | 0.007127 | 2.59.E-05 | 1.35 |
| *KMT2D* S696_D704del | rs780334086 | 0.0160 | 0.0162 | 0.008253 | 3.22.E-02 | 0.98 |
| *NOTCH3* R10_R13del | rs1219362463 | 0.0109 | 0.0119 | 0.005967 | 2.63.E-02 | 0.92 |
| *GNAS* P453_D464del | rs1196950915 | 0.0092 | 0.0337 | 0.01701 | 2.57.E-57 | 0.27 |
| *TSC2* D1690fs*27 † | rs778518656 | 0.0045 | 0.00015 | 0.000074 | 7.09.E-31 | 30.50 |
| *MTOR* T1830_T1833del | rs571156267 | 0.0039 | 0.0024 | 0.001225 | 2.93.E-03 | 1.58 |

† The variant was included because the allele frequency of another database, ALFA_Asian, was 0.0357, higher than that of ToMMo.

* ToMMo 54K_JPN database

### Table S3

Table S3. Frequency of variants classified by VAF category in the cases with increased somatic fraction (ISF) or associated conditions

| Category | Total (11,399 patients) | | Cases with high-VAF rPPSV (325) | Cases without high-VAF rPPSV (11,074) | Cases with C2, 4, and 5 rare autosomal variant(s) (2,589) | | Cases with ISF (2,602) | | Cases without ISF (8,797) | |
|---|---|---|---|---|---|---|---|---|---|---|
| | Number of variants | Number of rare autosomal variants | Number of rare autosomal variants, other than 341 high-VAF rPPSVs (per case) | Number of rare autosomal variants (per case) | Number of variants | Number of rare autosomal variants (per case) | Number of variants | Number of rare autosomal variants (per case) | Number of variants | Number of rare autosomal variants (per case) |
| C1 | 100,976 | 96,023 | 4,430 (13.63) | 91,593 (8.27) | 35,390 | 33,353 (12.88) | 35,506 | 33,458 (12.86) | 65,470 | 62,565 (7.11) |
| C2 | 6,923 | 5,328 | 805 (2.48) | 4,523 (0.41) | 6,505 | 5,328 (2.11) | 6,512 | 5,328 (2.05) | 411 | 0 (0.00) |
| C3 | 57,787 | 26,580 | 734 (2.26) | 25,846 (2.33) | 11,753 | 5,808 (2.29) | 11,836 | 5,849 (2.25) | 45,951 | 20,731 (2.36) |
| C4 | 2,307 | 1,259 | 232 (0.71) | 1,027 (0.09) | 2,096 | 1,259 (0.50) | 2,100 | 1,259 (0.50) | 207 | 0 (0.00) |
| C5 | 565 | 287 | 56 (0.17) | 231 (0.02) | 475 | 287 (0.11) | 475 | 287 (0.11) | 90 | 0 (0.00) |
| C6 | 812 | 76 | 2 (0.01) | 74 (0.01) | 251 | 76 (0.03) | 251 | 76 (0.03) | 561 | 0 (0.00) |

**Table S4. Criteria for variant detection numbers by category identified in each case to extract cases with high-VAF rPPSVs.**

| VAF Category | AUC (95% C.I.) | Cut-off value (variant count) | Sensitivity | Specificity |
|---|---|---|---|---|
| C1 | 0.602 (0.569-0.636) | 16 | 0.246 | 0.911 |
| C2 | 0.831 (0.808-0.854) | 1 | 0.806 | 0.823 |
| C3 | 0.477 (0.446-0.508) | 10 | 0.009 | 0.999 |
| C4 | 0.689 (0.661-0.716) | 1 | 0.440 | 0.933 |
| C5 | 0.553 (0.535-0.570) | 1 | 0.120 | 0.986 |
| C2+1 | 0.681 (0.652-0.710) | 9 | 0.606 | 0.659 |
| C2+3 | 0.705 (0.675-0.735) | 4 | 0.594 | 0.717 |
| C2+4 | 0.871 (0.851-0.890) | 1 | 0.883 | 0.802 |
| C2+5 | 0.841 (0.818-0.863) | 1 | 0.825 | 0.819 |
| C1+2+3 | 0.666 (0.636-0.697) | 11 | 0.622 | 0.626 |
| C1+2+4 | 0.706 (0.678-0.734) | 9 | 0.646 | 0.654 |
| C1+2+5 | 0.686 (0.657-0.715) | 9 | 0.609 | 0.658 |
| C2+3+4 | 0.764 (0.736-0.791) | 4 | 0.720 | 0.701 |
| C2+3+5 | 0.722 (0.693-0.752) | 4 | 0.631 | 0.713 |
| **C2+4+5** | **0.874 (0.855-0.894)** | **1** | **0.889** | **0.799** |
| C2+3+4+5 | 0.776 (0.749-0.802) | 4 | 0.742 | 0.697 |
| C1+2+4+5 | 0.711 (0.684-0.738) | 9 | 0.649 | 0.652 |
| C1+2+3+5 | 0.671 (0.641-0.701) | 11 | 0.628 | 0.625 |
| C1+2+3+4 | 0.691 (0.662-0.720) | 13 | 0.529 | 0.735 |
| All | 0.696 (0.667-0.724) | 11 | 0.646 | 0.620 |

### Table S5

**Table S5. Cases where pathogenic variants in genes with low or uncertain germline conversion rate (GCR) were identified in association with increased somatic fraction (ISF)**

| Research case # | Gender | Age | Gene symbol | Variant | VAF | Cancer type classification (OncoTree code) † | Family History † | Other pathogenic variants, used to determine ISF |
|---|---|---|---|---|---|---|---|---|
| 9317 | F | 70's | HNF1A | P291fs*51 | 0.32 | Adenocarcinoma, NOS (ADNOS) | None | KRAS G12D (0.35), TP53 Q144* (0.2), PIK3C2B C561fs*18 (0.2) |
| 3259 | M | 70's | NF2 | E317* | 0.3097 | Pancreatic Adenocarcinoma (PAAD) | Parent Stomach pa. 60's; Parent Pancreatic ca. ?'s; Parent Stomach ca. 70's | ATM c.72+2T>A (0.5841),<br>KRAS G12D (0.4793), ARID1A Y551fs*68 (0.3602) |
| 6464 | F | 80's | NF2 | W184* | 0.4117 | Salivary Carcinoma (SACA) | Child Breast ca. 40's; TDR** Uterine endometorioid ca. 40's | TP53 R273L (0.4701),<br>KMT2D Q1402* (0.2108) |
| 8387 | M | 50's | NF2 | c.599+1G>A | 0.49 | Lung Adenosquamous Carcinoma (LUAS) | None | EGFR E746_A750del (0.66), TP53 H179R (0.46) |
| 199 | M | 70's | PTCH1 | R1308fs*64 | 0.4241 | Prostate Adenocarcinoma (PRAD) | SDR* Breast ca. 20's | MSH3 K383fs*32 (0.3846), RB1 R552* (0.3637),<br>PALB2 N442fs*10 (0.3628), GNAS R201H (0.3405),<br>ASXL1 G646fs*12 (0.3069), CTCF E363fs*30 (0.215) |
| 84 | F | 60's | SMAD4 | R361H | 0.3529 | Pancreatic Adenocarcinoma (PAAD) | Sibling Kidney ca. 60's | KRAS G12D (0.4622), KMT2D S510* (0.3018), TP53 T125T (0.288) |
| 120 | M | 70's | SMAD4 | E330K | 0.4713 | Gallbladder Cancer (GBC) | Sibling Unknown ca. ?y | TP53 R326* (0.4522) |
| 818 | F | 60's | SMAD4 | I314fs*8 | 0.3883 | Gallbladder Adenocarcinoma, NOS (GBAD) | Parent Unknown ca. ?y; TDR** Unknown ca. ?y | NF1 E1929* (0.3998), TP53 R273H (0.3963),<br>PIK3CA E542K (0.2972), ASXL1 R625* (0.2504) |
| 832 | M | 70's | SMAD4 | L536fs*38 | 0.3276 | Cholangiocarcinoma (CHOL) | Unknown | KDM6A W1275* (0.4605), PIK3C2B c.2678+1G>A (0.431), RB1 L171fs*8 (0.4276),<br>PIK3CA C420R (0.288) |
| 1739 | F | 60's | SMAD4 | G352V | 0.3154 | **Colon Adenocarcinoma (COAD)** | Parent Prostate ca. 70's; SDR* Pharyngeal ca. 60's | TP53 A86fs*63 (0.3343), KRAS G12V (0.2969), AMER1 E597* (0.2724) |
| 1746 | M | 70's | SMAD4 | Q366* | 0.4264 | Colorectal Adenocarcinoma (COADREAD) | None | APC E225* (0.5119), ARID1A Q670* (0.3097), SOX9 S431fs*147 (0.2841) |
| 2264 | F | 60's | SMAD4 | W323* | 0.3049 | Pancreatic Adenocarcinoma (PAAD) | SDR* Stomach ca. 60's | BCL2L2 L13fs*13 (0.5154), TP53 R213* (0.4079), KRAS G12D (0.3728),<br>ARID1A Q520* (0.2167), ARID1A N116fs*11 (0.2099) |
| 2463 | M | 80's | SMAD4 | W302* | 0.407 | **Rectal Adenocarcinoma (READ)** | None | APC Y935fs*1 (0.4393), TP53 C124fs*25 (0.4368),<br>KRAS G12D (0.4191), PTEN L247* (0.414) |
| 2474 | F | 60's | SMAD4 | L238fs*66 | 0.5054 | **Rectal Adenocarcinoma (READ)** | Unknown | APC E1309* (0.784), TP53 G244C (0.7716),<br>KRAS Q61L (0.5489) |
| 2742 | M | 70's | SMAD4 | G386R | 0.5486 | Stomach Adenocarcinoma (STAD) | None | KRAS G12D (0.5971), APC R1450* (0.4493),<br>TP53 E286K (0.3705), TP53 R248Q (0.3009),<br>APC E1155fs*7 (0.2103) |
| 2850 | M | 70's | SMAD4 | R361H | 0.8564 | Gallbladder Cancer (GBC) | Parent Oral cavity ca. 60's | ARID1A D1689fs*11 (0.8783), TP53 D281Y (0.8228) |
| 3182 | M | 70's | SMAD4 | G168* | 0.42 | Intrahepatic Cholangiocarcinoma (IHCH) | Sibling Esophageal ca. 70's; Sibling Pharyngeal ca. 70's | KRAS G12C (0.7812), TP53 C275F (0.3967) |
| 4010 | M | 50's | SMAD4 | R445* | 0.6581 | **Colon Adenocarcinoma (COAD)** | Unknown | TP53 R196* (0.6612) |
| 4981 | F | 70's | SMAD4 | Q442* | 0.3156 | **Rectal Adenocarcinoma (READ)** | None | TP53 R175H (0.4254), APC T1556fs*3 (0.2124),<br>APC T1160fs*5 (0.2023) |
| 4989 | M | 60's | SMAD4 | R361H | 0.4815 | **Colon Adenocarcinoma (COAD)** | Unknown | TP53 R282W (0.4392), KRAS G12D (0.4181), SOX9 Q412* (0.3723),<br>APC R876* (0.2855), APC S1503* (0.2063) |
| 5503 | M | 60's | SMAD4 | R361P | 0.4484 | Gallbladder Cancer (GBC) | Parent Stomach 80's; Parent Lymphatic tumor 70's; SDR* Pharyngeal ca. 60's; SDR* Liver ca. 70's | ARID1A M954fs*47 (0.3742), APC T683fs*33 (0.286) |
| 6085 | M | 60's | SMAD4 | R361G | 0.59 | Non-Seminomatous Germ Cell Tumor (NSGCT) | Parent Pancreatic ca. 80's; Parent Stomach ca. 60's; SDR* Pancreatic ca. >90y | ARID1A E2250fs*28 (0.56), TP53 c.376-20_422del (0.37) |
| 6139 | M | 70's | SMAD4 | S357fs*20 | 0.38 | Pancreatic Adenocarcinoma (PAAD) | Sibling Liver ca. 60's | BRCA2 R2318* (0.69), TP53 L194R (0.39),<br>KRAS G12D (0.27) |
| 6402 | M | 50's | SMAD4 | G89* | 0.3805 | **Colon Adenocarcinoma (COAD)** | None | APC E1379* (0.512), TP53 H193R (0.493), SOX9 Q376* (0.4244), BRAF D594G (0.4062), FBXW7 c.986-1G>A (0.2167) |
| 6631 | F | 40's | SMAD4 | R361S | 0.6959 | **Colon Adenocarcinoma (COAD)** | Sibling Breast ca. 30's | TP53 R248W (0.6706), BRAF V600E (0.514),<br>APC T1556fs*3 (0.447), APC E1132* (0.3503) |
| 7317 | F | 40's | SMAD4 | R361C | 0.62 | **Colon Adenocarcinoma (COAD)** | SDR* Unknown ca. 60's; SDR* Lung ca. 70's | APC L1506fs*1 (0.65), KRAS Q61K (0.58),<br>TP53 R273H (0.55) |
| 7522 | M | 60's | SMAD4 | R361H | 0.43 | **Colon Adenocarcinoma (COAD)** | Parent Liver ca. 70's; Sibling Breast ca. 40's; TDR** Menigeal tumor 50's | TP53 R175H (0.47), KRAS G12V (0.34), APC R564* (0.29), APC S1400fs*1 (0.27),<br>SOX9 Q339* (0.26) |
| 7742 | F | 60's | SMAD4 | G231fs*10 | 0.57 | **Rectal Adenocarcinoma (READ)** | Parent Lung ca. ?y | APC A1316fs*5 (0.64), KEL E239* (0.5), TP53 R175H (0.48), PIK3CA R38H (0.44) |
| 7850 | M | 80's | SMAD4 | R361C | 0.59 | **Colon Adenocarcinoma (COAD)** | Parent Unknown ca. ?y; Sibling Pancreatic ca. 70's | TP53 Y220C (0.74), AMER1 F536fs*6 (0.69), KRAS G13D (0.53), APC R1450* (0.4),<br>APC L508* (0.33) |
| 8379 | M | 40's | SMAD4 | D351V | 0.43 | **Rectal Adenocarcinoma (READ)** | Parent Stomach ca. ?y; Parent Colon ca. ?y; SDR* Stomach ca. ?y; SDR* Oral cavity ca. ?y; | APC S1355fs*1 (0.22) |
| 8495 | M | 50's | SMAD4 | E330K | 0.33 | Pancreatic Adenocarcinoma (PAAD) | None | KRAS G12D (0.56), TP53 R248Q (0.38) |
| 8927 | M | 50's | SMAD4 | Q461fs*15 | 0.62 | Pancreatic Adenocarcinoma (PAAD) | Parent Lung ca. 60's | KRAS G12R (0.67), TP53 C242fs*5 (0.61) |
| 9079 | M | 60's | SMAD4 | L146fs*9 | 0.3 | Pancreatic Adenocarcinoma (PAAD) | Parent Esocphageal ca. 60's; SDR* Pancreatic ca. 60's; TDR** Lung ca. 60's | KDM6A K1076fs*5 (0.4), KRAS G12D (0.29) |
| 9468 | F | 60's | SMAD4 | R361H | 0.56 | Intrahepatic Cholangiocarcinoma (IHCH) | Parent Lymphatic tumor 80's; SDR* Myeloid tumor 60's; SDR* Stomach ca. 50's; SDR* Lung ca. 80's | ACVR1B L377fs*1 (0.5), TP53 E286* (0.45),<br>ARID1A Q790* (0.22) |
| 9601 | M | 70's | SMAD4 | A421fs*7 | 0.37 | Pancreatic Adenocarcinoma (PAAD) | None | KRAS G12D (0.4), TP53 P151T (0.38), KMD6A K716* (0.37) |
| 9722 | F | 60's | SMAD4 | R361H | 0.55 | **Colon Adenocarcinoma (COAD)** | Parent Unknown ca. 80's; SDR* Colon ca. 80's; SDR* Colon ca. 80's; SDR* Colon ca. 80's | TP53 R306* (0.52), KRAS G12V (0.5),<br>APC T1556fs*3 (0.28), APC R876* (0.27) |
| 9987 | M | 60's | SMAD4 | R361C | 0.47 | **Rectal Adenocarcinoma (READ)** | None | APC E1309fs*4 (0.59), TP53 R175H (0.45), FBXW7 R367* (0.28),<br>KRAS G12C (0.27) |
| 10535 | M | 60's | SMAD4 | Q334* | 0.49 | **Rectal Adenocarcinoma (READ)** | SDR* Lung ca. ?y | APC c.835-1G>A (0.67), TP53 E286K (0.65),<br>KRAS G12D (0.65) |
| 11272 | F | 40's | SMAD4 | R361G | 0.75 | **Colon Adenocarcinoma (COAD)** | Sibling Unknown ca. ?y; SDR* Unknown ca. ?y; SDR* Unknown ca. ?y | TP53 c.375+2T>G (0.76), APC c.835-8A>G (0.54),<br>KRAS G12D (0.53), APC V704fs*2 (0.28) |
| 366 | F | 60's | STK11<br>TGFBR2 | P203fs*84<br>E82* | 0.5672<br>0.6563 | Cervical Squamous Cell Carcinoma (CESC) | None | FBXW7 L660fs*34 (0.6715), TBX3 S214* (0.2614) |
| 1811 | F | 50's | STK11 | Q220* | 0.657 | Lung Adenocarcinoma (LUAD) | Parent Stomach ca. 40's; SDR* Lung ca. 80's; Sibling Cervical ca. 40's | TP53 P278R (0.6616), EGFR E746_T751delinsVA (0.4995) |
| 1868 | M | 70's | STK11 | I322fs*33 | 0.3037 | Intrahepatic Cholangiocarcinoma (IHCH) | Parent Lung ca. 80's | BCORL1 P1681fs*20 (0.7983), ARID1A P1898fs*25 (0.6556), PBRM1 Q478* (0.654),<br>BCL6 P473fs*11 (0.6219), APC F801fs*19 (0.618),<br>CDK12 c.1056+2T>C (0.4085), BRD4 P46fs*47 (0.4074), KDM5A G1200fs*9 (0.3986),<br>GNAS R201H (0.3933), CDH1 R63* (0.3875), PRKAR1A R96* (0.3778), ATR R1814fs*10 (0.3677), MRE11 T408fs*49 (0.2754) |
| 6451 | M | 70's | SMAD4<br>STK11 | V407fs*23<br>D53fs*110 | 0.3662<br>0.3255 | Intrahepatic Cholangiocarcinoma (IHCH) | None | RNF43 V31fs*9 (0.2842), CHEK2 Y220* (0.2677),<br>PIK3C2G Q326fs*7 (0.2571), NF1 I417fs*56 (0.2304) |
| 8602 | M | 60's | STK11 | K84* | 0.31 | Gallbladder Adenocarcinoma, NOS (GBAD) | Parent Stomach ca. 80's; SDR* Skin ca. ?y | ARID1A Q507* (0.22), KEAP1 Y584fs*1 (0.2) |
| 9318 | M | 60's | STK11 | D194G | 0.71 | **Pancreatic Adenocarcinoma (PAAD)** | None | APC E1416fs*8 (0.56), ARID1A S2262* (0.51), PIK3CA Y1021H (0.25) |
| 9481 | M | 70's | STK11 | D194N | 0.75 | **Rectal Adenocarcinoma (READ)** | Parent Colon Co. 70's | ATM R3047* (0.73), KRAS G13D (0.39), APC R564* (0.38), APC E1309fs*4 (0.32) |
| 3539 | M | 60's | TGFBR2<br>TGFBR2 | R528H<br>R528C | 0.3152<br>0.3195 | Pancreatic Adenocarcinoma (PAAD) | None | KDM5C V1036fs*2 (0.7223), TP53 R175H (0.5195),<br>KRAS G12D (0.332), KMT2D E1594fs*8 (0.3309) |
| 8031 | M | 60's | TGFBR2 | R528H | 0.37 | Colon Adenocarcinoma (COAD) | Parent Esophageal ca. 60's; SDR* CUP 60's; SDR* Lung ca. 50's | TP53 G245S (0.39), KRAS G13D (0.33), APC Q1067* (0.31) |
| 8982 | M | 50's | TGFBR2 | Q194* | 0.5 | Oropharynx Squamous Cell Carcinoma (OPHSC) | SDR* Unknown ca. ?y | TP53 P82fs*41 (0.51), PIK3CA E545K (0.28) |

* Second-degree relative, ** Third-degree relative, † Bold and undelined cancer types are associated with the concerned variants

### Table S6

**Table S6. Cases with presumed somatic and germline pathogenic variants with 0.2≤VAF<0.3 in high germline conversion rate genes**

| Research case # | Gene Symbol | Variant | Chr | Position (hg38) | Ref | Alt | VAF | Gender | Age at registration | Disease | Family history | Increased Somatic Fraction* | Other PGPVs | Putative reversion mutation in *BRCA1/2* |
|---|---|---|---|---|---|---|---|---|---|---|---|---|---|---|
| 3276 | *POLE* | S297F | 12 | 132676565 | G | A | 0.2977 | M | 70's | **Prostate Adenocarcinoma** | Sibling Esophageal ca. 70's | Yes | No | N/A |
| 7315 | *BRCA1*<br>*MSH2* | K654fs*47<br>E852* | 17<br>2 | 43093569<br>47480791 | CT<br>G | C<br>T | 0.29<br>0.29 | M | 70's | **Prostate Adenocarcinoma** | None | Yes | *MSH2*:p.Q374* (0.52) | N/A |
| 10180 | *BRCA2* | T3033fs*29 | 13 | 32379892 | AA | A | 0.29 | M | 70's | **Prostate Adenocarcinoma** | Sibling Prostate ca. 70's; Sibling Prostate ca. 70's | Yes | No | No |
| 3110 | *MSH6* | F1088fs*5 | 2 | 47803508 | C | CC | 0.2874 | M | 70's | Prostate Adenocarcinoma | None | Yes | *MSH6*:p.R1331* (0.3463) | N/A |
| 1699 | *ATM* | R3008C | 11 | 108365359 | C | T | 0.2861 | M | 70's | Gallbladder Cancer | None | No | No | N/A |
| 11347 | *ATM* | N3044fs*31 | 11 | 108365467 | AA | A | 0.28 | M | 60's | Prostate Adenocarcinoma | None | Yes | No | N/A |
| 437 | *BRIP1* | Y313fs*6 | 17 | 61801455 | T | TA | 0.2762 | M | 80's | Prostate Adenocarcinoma | Parent Prostate ca. 70's; Parent Myeloid tumor 70's | Yes | No | N/A |
| 5821 | *ATM* | R2993* | 11 | 108365208 | C | T | 0.2758 | M | 70's | Stomach Adenocarcinoma | Parent Colon ca. 60's; Parent Stomach ca. 70's; SDR* Colon ca. 60's; Sibling Prostate ca. 70's | Yes | No | N/A |
| 352 | *BRCA1* | c.5075-1G>C | 17 | 43063952 | C | G | 0.2727 | F | 50's | Gallbladder Adenocarcinoma, NOS | SDR* Colon ca. ?y | Yes | No | No |
| 1630 | *BRCA2* | E804* | 13 | 32336765 | G | T | 0.2705 | M | 50's | Intrahepatic Cholangiocarcinoma | Unknown | Yes | *MLH1*:c.545+1G>T (0.4094) | No |
| 9765 | *BRCA2* | S3080fs*3 | 13 | 32380126 | TT | T | 0.27 | M | 70's | **Prostate Adenocarcinoma** | None | Yes | No | No |
| 8464 | *MLH1* | R182G | 3 | 37008904 | A | G | 0.27 | M | 60's | **Poorly Differentiated Carcinoma of the Stomach** | SDR* Pancreatic ca. 80's; Parent Myeloid tumor 70's; TDR** Stomach ca. 60's; SDR* Brain tumor 50's | Yes | No | N/A |
| 3025 | *ATM* | c.7308-1G>C | 11 | 108330213 | G | C | 0.2688 | M | 70's | **Prostate Adenocarcinoma** | None | No | No | N/A |
| 4078 | *MSH2* | V257fs*24 | 2 | 47412535 | CTGTAT TGC | C | 0.2685 | M | 40's | Adenocarcinoma, NOS | SDR* Breast ca. ?y; SDR* Breast ca. ?y; SDR* Pancreatic ca. ?y | Yes | No | N/A |
| 6451 | *CHEK2* | Y220* | 22 | 28719418 | G | GTAT | 0.2677 | M | 70's | Intrahepatic Cholangiocarcinoma | None | Yes | No | N/A |
| 4115 | *ATM* | c.663-2A>G | 11 | 108244786 | A | G | 0.2663 | M | 60's | Well-Differentiated Neuroendocrine Tumor of the Rectum | Sibling Breast ca. 60's; Parent Pancreatic ca. 80's; Parent Esophageal ca. ?y; SDR* Ovary / fallopian tube ca. 50's; SDR* Prostate ca. ?y | Yes | No | N/A |
| 4503 | *FLCN* | G195fs*28 | 17 | 17223955 | TC | T | 0.261 | M | 60's | Lung Adenocarcinoma | Parent Stomach ca. 70's; SDR* Lung ca. 70's | Yes | No | N/A |
| 9403 | *ATM* | c.4909+2T>C | 11 | 108295061 | T | C | 0.26 | M | 80's | **Prostate Adenocarcinoma** | Parent Prostate ca. 80's; SDR* Brain tumor <10y; SDR* Stomach ca. 30's | No | No | N/A |
| 10019 | *ATM* | M2531fs*5 | 11 | 108331519 | AT | A | 0.26 | M | 70's | **Prostate Adenocarcinoma** | SDR* Unknown ca. 70's | No | No | N/A |
| 7523 | *BRCA2* | E1717* | 13 | 32339504 | G | T | 0.26 | F | 50's | **Pancreatic Adenocarcinoma** | Parent Breast ca. ?y; Sibling Lymphatic tumor ?y; SDR* Breast ca. ?y; SDR* Breast ca. ?y; SDR* Pancreatic ca. ?y; SDR* Prostate ca. ?y | Yes | *BRCA2*:p.M815fs*10 (0.46) | *BRCA2*:p.M815_L824delinsWKRIKMYV (0.13), *BRCA2*:p,K811_P814del (0.01), *BRCA2*:p.1196_L2240del (0) |
| 9245 | *FLCN* | H429fs*27 | 17 | 17216394 | T | TG | 0.26 | M | 70's | Prostate Adenocarcinoma | None | Yes | No | N/A |
| 8714 | *MSH2* | R389* | 2 | 47429830 | C | T | 0.26 | F | 70's | **Gallbladder Adenocarcinoma, NOS** | Parent Breast ca. 70's; Sibling Lung ca. 70's | No | No | N/A |
| 10892 | *MSH6* | Q618* | 2 | 47799835 | C | T | 0.26 | F | 50's | **Intrahepatic Cholangiocarcinoma** | None | Yes | No | N/A |
| 5676 | *ATM* | Y1844* | 11 | 108304710 | C | A | 0.2577 | F | 40's | Rectal Adenocarcinoma | None | Yes | No | N/A |
| 5245 | *MLH1*<br>*BRCA1*<br>*MSH6*<br>*FLCN* | A681T<br>c.5468-1G>A<br>F1088fs*5<br>H429fs*27 | 3<br>17<br>2<br>17 | 37048955<br>43045803<br>47803508<br>17216394 | G<br>C<br>C<br>T | A<br>T<br>CC<br>TG | 0.2507<br>0.2399<br>0.2346<br>0.2013 | M | 70's | **Prostate Adenocarcinoma** | SDR* Lung ca. 70's | Yes | *MSH2*:p.E878* (0.3348) | No |
| 6917 | *BRCA2* | D946fs*13 | 13 | 32337190 | A | AA | 0.2494 | F | 50's | **Breast Invasive Ductal Carcinoma** | Parent Bile duct ca. 50's; SDR* Breast ca. 30's; SDR* Breast ca. 70's; TDR* CUP# 40's; TDR* Liver ca. 40's; TDR* CUP# 40's | Yes | *BRCA2*:p.A938fs*21 (0.6464) | *BRCA2*:p.S942fs*2 (0.0267), *BRCA2*:p.Q940fs*2 (0.0145), *BRCA2*:p.S942fs*3 (0.001), etc |
| 665 | *BRIP1* | I552_Q553delinsN* | 17 | 61780977 | GAA | AAT | 0.2432 | F | 60's | **Invasive Breast Carcinoma** | Unknown | Yes | No | N/A |
| 2061 | *BRCA2* | Q2163* | 13 | 32340842 | C | T | 0.2421 | M | 50's | **Prostate Adenocarcinoma** | Parent Liver ca 70's | Yes | No | No |
| 9635 | *BRCA1* | E1731* | 17 | 43063335 | C | A | 0.24 | F | 60's | Bladder Urothelial Carcinoma | Parent Stomach ca. 70's; Parent Colon ca. 90's; SDR* Laryngeal ca. 60's; SDR* Stomach ca. 60's | Yes | No | No |
| 5198 | *BRCA2* | I1859fs*3 | 13 | 32339930 | ATTAA | A | 0.2382 | M | 70's | **Prostate Adenocarcinoma** | Sibling Bile duct ca. 70's; Daughter Endmetrioid ca. 40's; TDR* Liver ca. 50's | Yes | No | *BRCA2*:p.S1650_K1888del (0.0061); *BRCA2*:p.A1847_I1859del (0.0054), etc |
| 9188 | *MLH1* | I611fs*3 | 3 | 37047618 | A | AA | 0.23 | M | 60's | **Intrahepatic Cholangiocarcinoma** | Parent Colon ca. 70's; SDR* Stomach ca. ?y | Yes | No | N/A |
| 9270 | *PMS2* | I143fs*58 | 7 | 6002566 | TC | T | 0.23 | M | 60's | Undifferentiated Malignant Neoplasm | Unknown | Yes | No | N/A |
| 4731 | *BRCA2* | P704fs*26 | 13 | 32336465 | CC | C | 0.2277 | M | 70's | **Prostate Adenocarcinoma** | Unknown | Yes | *BRCA2*:p.I1859fs*3 (0.4825) | No |
| 3432 | *ATM* | I1422fs*29 | 11 | 108289627 | CC | C | 0.2223 | M | 70's | **Pancreatic Adenocarcinoma** | Parent Liver ca. 70's; Parent Pancreatic ca. 70's | Yes | No | N/A |
| 1550 | *MLH1* | E717* | 3 | 37050531 | G | T | 0.2209 | F | 50's | Breast Invasive Ductal Carcinoma | Parent Prostate ca. 70's | Yes | No | N/A |
| 8257 | *MSH6* | F1088fs*5 | 2 | 47803508 | C | CC | 0.22 | M | 60's | **Colon Adenocarcinoma** | SDR* Pancreatic ca. 60's; SDR* Colon ca. 90's; TDR* Colon ca. 50's | Yes | No | N/A |
| 1585 | *ATM* | D2708N | 11 | 108335080 | G | A | 0.2191 | M | 60's | Lung Adenocarcinoma | Sibling Lung ca. 40's; Parent Breast ca. 80's | No | *ATM*:p.R1973fs*2 (0.2675) † | N/A |
| 5463 | *BRCA1* | K654fs*47 | 17 | 43093569 | CT | C | 0.2179 | F | 40's | Uterine Serous Carcinoma/Uterine Papillary Serous Carcinoma | SDR* Breast ca. 40's; SDR* Prostate ca. 70's; Parent Thyroid ca. 60's | Yes | No | No |
| 4308 | *CHEK2* | A98fs*10 | 22 | 28734423 | TGAAG GGC | T | 0.2156 | F | 80's | Pancreatic Adenocarcinoma | Sibling Unknown ca. ?y; Sibling Unknown ca. ?y | Yes | No | N/A |
| 5065 | *FLCN* | H429fs*39 | 17 | 17216394 | TG | T | 0.2113 | F | 60's | Breast Invasive Ductal Carcinoma | None | Yes | No | N/A |
| 10006 | *ATM* | c.3078-1G>T | 11 | 108272531 | G | T | 0.21 | M | 50's | Esophageal Squamous Cell Carcinoma | SDR* Lung ca. 70's; SDR* Lung ca. 80's; SDR* Colon ca. 70's | Yes | No | N/A |
| 11008 | *ATM* | E2837* | 11 | 108345833 | G | T | 0.21 | M | 60's | Prostate Adenocarcinoma | Unknown | No | No | N/A |
| 2188 | *BRIP1* | L477fs*34 | 17 | 61793639 | T | TA | 0.21 | M | 70's | Small Cell Carcinoma of Unknown Primary | Parent Lymphatic tumor 60's | Yes | No | N/A |
| 10197 | *MSH6* | F1088fs*2 | 2 | 47803507 | CC | C | 0.21 | M | 30's | **Intrahepatic Cholangiocarcinoma** | SDR* Oral cavity ca. 70's; SDR* Myeloid tumor 70's; SDR* Bile duct ca. 70's; SDR* Lung ca. 70's | Yes | *MLH1*:p.L650fs*35 (0.41) | N/A |
| 573 | *ATM* | R2832C | 11 | 108345818 | C | T | 0.2073 | M | 60's | Colon Adenocarcinoma | Unknown | Yes | No | N/A |
| 3590 | *BRCA2* | P3039P | 13 | 32379913 | G | A | 0.2057 | M | 70's | **Prostate Adenocarcinoma** | None | No | No | No |
| 5691 | *ATM* | R250* | 11 | 108244873 | C | T | 0.2038 | M | 70's | Perihilar Cholangiocarcinoma | Parent Prostate ca. ?y | No | No | N/A |
| 2969 | *ATM* | c.2124+1G>A | 11 | 108254040 | G | A | 0.2022 | M | 70's | Gallbladder Adenocarcinoma, NOS | None | Yes | No | N/A |
| 8088 | *BRCA2* | S1099fs*1 | 13 | 32337650 | TC | T | 0.2 | M | 60's | **Prostate Adenocarcinoma** | Parent Kidney ca. ?y; Parent Stomach ca ?y; Parent Liver ca. ?y; SDR* Breast ca. ?y | Yes | No | No |
| 9475 | *BRIP1* | Q1010* | 17 | 61684018 | G | A | 0.2 | F | 70's | Extrahepatic Cholangiocarcinoma | Parent Liver ca. 60's; Sibling Colon ca. 70's; SDR* Stomach ca. 70's | Yes | No | N/A |

Case #5198, in red, harbored somatic reversion mutations of presumed germline pathogenic variants, *BRCA2* p.I1859fs*3 (VAF 0.2382). * Second-degree relative, ** Third-degree relative, # Cancer of unknown primary, † ATM:p.R1973fs*2 is described in **Table S7** (Case#1585).

### Table S7

**Table S7. Cases with presumed germline pathogenic variants with long indel and 0.2≤VAF<0.3**

| Research Case # | Gene Symbol | Variant | Chr | Position (hg38) | Ref | Alt | indel length (bp) | VAF | Gender | Age at registration | Disease | Family history |
|---|---|---|---|---|---|---|---|---|---|---|---|---|
| 1011 | *ATM* | Y1415fs*5 | 11 | 108,289,608_ 108,289,622 | TATCAGAAAATTCTT | T | 14 | 0.2077 | M | 60's | Lung Adenocarcinoma | Sibling Oral cavity ca. 60's |
| 10493 | *BRCA1* | c.4085_4096+19del 31 | 17 | 43,091,415_ 43,091,446 | CAAAAACCTGGTTCCA ATACCTAAGTTTGAAT | C | 31 | 0.22 | M | 60's | **Prostate Adenocarcinoma** | None |
| 2797 | *MSH6* | I464fs*7 | 2 | 47,799,372_ 47,799,398 | AATTGCATTTGGCCGT TATTCAGATTC | A | 26 | 0.265 | F | 60's | Small Cell Bladder Cancer | Sibling **Endometrial ca.** ?y; SDR* **Colon ca.** ?y; SDR* Stomach ca. ?y; TDR** Colon ca. ?y |
| 1585 | *ATM* | R1973fs*2 | 11 | 108,312,410_ 108,312,421 | GAAGTCTTGCAT | G | 11 | 0.2675 | M | 60's | Lung Adenocarcinoma | Sibling Lung ca. 40's; Parent **Breast ca.** 80's |

* Second-degree relative, ** Third-degree relative

### Table S8

**Table S8. Cases with presumed germline pathogenic variants of TP53, selected by the GeMSort algorithm.**

| Research Case # | TP53 variant | | VAF | Gender | Age | Cancer type classification | Other genomic alterations | Other cancers (counts) | Multiple cancers from same organ | Family history |
|---|---|---|---|---|---|---|---|---|---|---|
| 130 | *TP53* | T312fs*24 | 0.8424 | M | 50's | Small Cell Lung Cancer (SCLC) | *HRAS* G13V (0.0021), *PIK3CA* E542K (0.0038), *AKT3* E17K (0.0044), *PIK3CA* N345K (0.0052), *ARID1A* D1850fs*33 (0.0119), *KDM6A* Q880fs*16 (0.0147), *PIK3CA* Q546K (0.0148), *PIK3CA* H1047L (0.0351) | None | None | Parent Unknown ca. ?y, Parent Unknown ca. ?y |
| 177 | *TP53* | R175H | 0.5836 | M | 70's | Lung Adenocarcinoma (LUAD) | *ERRFI1* S58fs*45 (0.0031) | **Yes (1)** | **Yes** | Unknown |
| 738 | *TP53* | V173L | 0.4984 | F | 40's | Breast Invasive Ductal Carcinoma (IDC) | No other driver mutations | **Yes (2)** | None | Parent Uterine ca. **30's**, Sibling Oral cavity ca. **20's**, SDR* Pancreatic ca ?y |
| 1975 | *TP53* | R181H | 0.5119 | F | 30's | Rectal Adenocarcinoma (READ) | *KRAS* G12D (0.0121), *PIK3CA* E545K (0.0125) | None | None | Parent Stomach ca. **20's**, SDR* Colon ca. 80's, SDR* Stomach ca. 70's, SDR* Oral cavity ca. 60's, TDR** Lymphatic tumor 50's |
| 2206 | *TP53* | R273H | 0.4443 | M | 40's | Pancreatic Adenocarcinoma (PAAD) | No other driver mutations | None | None | Parent Bile duct ca. 60's, SDR* Laryngeal ca. ?y |
| 2412 | *TP53* | G245S | 0.5045 | M | 20's | Intestinal Type Stomach Adenocarcinoma (ISTAD) | *CDKN2A* W15* (0.0053) | None | None | Parent Pancreatic ca. **30's**, Sibling Liver ca. **<10y**, SDR* Unknown ca. ?y |
| 3173 | *TP53* | R175H | 0.5238 | F | 50's | Pancreatic Adenocarcinoma (PAAD) | *KMT2D* Q3813* (0.0139), *CDKN2A* Q50* (0.014), *KRAS* G12V (0.0157) | **Yes (2)** | None | Parent Lung ca. 60's, SDR* Lung ca. ?y |
| 4817 | *TP53* | R158H | 0.5041 | M | 40's | **Leiomyosarcoma** (LMS) | *FANCA* S849fs*40 (0.4647) | None | None | Parent Stomach ca. ?y; Parent Breast ca. ?y, SDR* Lung ca. ?y, SDR* Unknown ca. ?y, SDR* Unknown ca. ?y |
| 5157 | *TP53* | A276P | 0.5323 | M | 50's | Prostate Adenocarcinoma (PRAD) | *CHEK1* E104fs*4 (0.0385) | None | None | Parent Stomach ca. 40's, Sibling **Brain tumor 10's**, SDR* Esophageal ca. 50's |
| 6350 | *TP53* | P27fs*17 | 0.5402 | M | 50's | Prostate Adenocarcinoma (PRAD) | No other driver mutations | None | None | None |
| 6642 | *TP53* | R175H | 0.5039 | F | 40's | Pancreatic Adenocarcinoma (PAAD) | No other driver mutations | **Yes (3)** | None | Parent Colon ca. ?y, Parent **Breast ca. 30's**, Parent Stomach ca. 40's, Sibling Lung ca. 40's, Sibling Breast ca. 40's, Sibling Other ca. ?y |
| 6752 | *TP53* | R248W | 0.5147 | F | 30's | Lung Adenocarcinoma (LUAD) | *EGFR* D770delinsNNNN (0.0044) | **Yes (1)** | None | SDR* Bladder ca. 70's, SDR* Colon ca. 70's |
| 7919 | *TP53* | C277Y | 0.41 | M | 10's | **Alveolar Rhabdomyosarcoma** (ARMS) | *TP53* P278L (0) | None | None | Parent Cervical ca. 40's, SDR* Thyroid ca. 60's |
| 9081 | *TP53* | R273C | 0.48 | M | <10y | **CNS/Brain** (BRAIN) | No other driver mutations | Unknown | Unknown | Unknown |
| 9374 | *TP53* | R333fs*12 | 0.52 | F | 40's | Lung Adenocarcinoma (LUAD) | *EGFR* L747_S752del (0.04) | None | None | Parent **Bone sarcoma 10's**, Child **Soft tisssue sa. <10y** |
| 9457 | *TP53* | S127F | 0.49 | F | 60's | Inflammatory Breast Cancer (IBC) | *TET2* L615fs*24 (0.01) | None | **Yes** | Sibling Stomach ca. 40's, Sibling Colon ca. 50's, Parent Lung ca. 70's |
| 9499 | *TP53* | R196* | 0.48 | F | 20's | **Diffuse Astrocytoma** (DASTR) | No other driver mutations | None | None | Sibling Colon ca. **20's** |

* Second-degree relative, ** Third-degree relative
